# Projecting climate change impacts on inter-epidemic risk of Rift Valley fever across East Africa

**DOI:** 10.1101/2025.02.08.25321747

**Authors:** Evan A. Eskew, Erin Clancey, Deepti Singh, Silvia Situma, Luke Nyakarahuka, M. Kariuki Njenga, Scott L. Nuismer

**Affiliations:** Institute for Interdisciplinary Data Sciences, University of Idaho, Moscow ID, USA; Paul G. Allen School for Global Health, Washington State University, Pullman WA, USA; School of the Environment, Washington State University, Vancouver WA, USA; Washington State University Global Health-Kenya, Nairobi, Kenya; Department of Animal Science, Pwani University, Kilifi, Kenya; Department of Biosecurity, Ecosystems & Veterinary Public Health, Makerere University, Kampala, Uganda; Uganda Virus Research Institute, Entebbe, Uganda; Department of Biological Sciences, University of Idaho, Moscow ID, USA

**Keywords:** Coupled Model Intercomparison Project, emerging infectious disease, machine learning, shared socioeconomic pathways, vector-borne disease, zoonosis

## Abstract

**Background:** Rift Valley fever (RVF) is a zoonotic disease that causes sporadic, multi-country epidemics. However, RVF virus (RVFV) also circulates during inter-epidemic periods. There is limited understanding of how climate change will affect inter-epidemic RVF. Here, we project inter-epidemic RVF risk under future climate scenarios, focusing on the East African countries of Kenya, Tanzania, and Uganda.

**Methods:** We combined data on inter-epidemic RVF outbreaks and spatially-explicit predictor variables to build a predictive model of inter-epidemic RVF risk. We validated our model using RVFV serological data from humans. We then projected inter-epidemic RVF risk for three future time periods (2021-2040, 2041-2060, 2061-208) under three climate scenarios (SSP126, SSP245, SSP370). Finally, we combined risk projections with human population projections to estimate the future population at risk of inter-epidemic RVF across the study region.

**Findings:** Our model showed seasonality in inter-epidemic RVF, with risk peaking May-July following the long rains (March-May). Projections for future climate scenarios suggested that disease risk will increase January-March, with the present-day hotspots of east Kenya, southeast Tanzania, and southwest Uganda remaining high-risk. By 2061-2080, > 117 million people in the study region may be at risk from inter-epidemic RVF, a fourfold increase relative to the historical (1970-2000) estimate of ∼25 million people.

**Interpretation:** Climate change will shift the inter-epidemic RVF risk landscape, with increasing short rains (October-December) driving increased risk January-March. Mitigating the future health impacts of RVF will require increased disease surveillance, prevention, and control effort in risk hotspots.

**Funding:** US National Institutes of Health.

**RESEARCH IN CONTEXT:** *Evidence before this study:* As for arboviruses generally, global climate change may shift the geographic distribution, timing, and severity of disease burden imposed by Rift Valley fever (RVF). To review the existing evidence for climate change impacts on RVF while adopting a specific geographic focus on East Africa, we searched Web of Science using the query string “ALL=(climate change) AND ALL=(Rift Valley fever) AND ALL=(‘East Africa’ OR Kenya OR Tanzania OR Uganda)”. Our search returned 39 results published between 2007 and 2024. We reviewed these contents to identify work with a substantive focus on understanding how future climate change will affect RVF (as opposed to investigations discussing any connection between climatic factors and RVF occurrence). We identified a total of ten papers that met this criterion: two were reviews, four analyzed mechanistic compartmental models, and four were empirical papers adopting other modeling and analysis methods. These studies highlighted the key role of precipitation and temperature in RVF epidemiology. Heavy precipitation is a well-known RVF driver given that rainfall can cause large amounts of surface water to become available for mosquito vector breeding. As such, expected wetting trends in East Africa under climate change scenarios could fuel increased frequency and severity of RVF. Temperature influences on RVF are underappreciated relative to precipitation, but climate-driven compartmental models emphasize that RVFV transmission is likely to be highest in areas that maintain optimal temperatures for mosquito development (∼22-26°C).

*Added value of this study:* Adopting a machine learning approach, we found that precipitation, goat density, soil silt, and elevation were among the most important predictors of inter-epidemic RVF. Our model, which was trained on monthly climatic data, showed seasonally-varying RVF risk, with risk peaking following the long rains season (March-May) and, to a lesser degree, following the short rains (October-December). We used our trained model, which was validated against serological data from humans, to project RVF risk for three future time periods (2021- 2040, 2041-2060, 2061-208) under three climate scenarios (SSP126, SSP245, SSP370) using 11 climate models from the Coupled Model Intercomparison Project (CMIP6). As a result, our multi-model projections of inter-epidemic RVF risk explicitly incorporate multiple sources of climate uncertainty. Projections suggested that RVF risk will increase January-March across the study region, particularly under high-emissions scenarios (i.e., SSP370). Although there are some anticipated changes in the spatial distribution of RVF risk, future risk hotspots largely mirror the present-day, with high risk in east Kenya, southeast Tanzania, and southwest Uganda.

*Implications of all the available evidence:* Precipitation, the major driver of RVF, shapes both the temporal and spatial patterning of RVF risk across East Africa. Projections of future RVF risk are also strongly influenced by precipitation, with projected increases in disease risk January-March arising from projected increases in short rains (October-December) precipitation under future climate scenarios. Accurate projections of future precipitation, including a better understanding of potential changes to climatic linkages like the El Niño-Southern Oscillation and Indian Ocean Dipole, will enable meaningful prediction of future RVF risk that can inform disease interventions. Greater consideration of population-level host immunity and climate adaptation behaviors in East Africa (i.e., changing livestock management practices) would also allow for more realistic RVF risk projections.

## INTRODUCTION

Rift Valley fever (RVF), a mosquito-borne zoonotic disease caused by RVF virus (RVFV), affects Africa and the Arabian peninsula.^1^ RVF is a unique threat because, in addition to causing human morbidity and mortality, disease outbreaks can impact the livestock trade that supports hundreds of millions of livelihoods throughout Africa.^1,2^ To counteract RVF’s devastating health and economic burden, models have been developed to predict when and where major regional epidemics could emerge. RVF outbreaks often occur following excessive rainfall^3^ when flooding creates aquatic breeding habitats that fuel the growth of RVFV-infected mosquito populations.^1,4,5^ Disease forecasts have leveraged this information by using the normalized difference vegetation index (NDVI), a satellite-derived measure of vegetation growth in response to rainfall, to predict impending RVF epidemics.^4,6^ This near-term forecasting approach enabled prediction of the multi-country 2006-2007 RVF epidemic weeks before it occurred, facilitating early warning of the at-risk human population.^7^

Despite the promise of these predictive methods, the traditional research focus on large, but relatively infrequent, regional RVF epidemics is challenged by increased recognition of RVF activity during inter-epidemic periods.^8,9^ For example, RVFV circulates endemically in humans across East Africa, including in Kenya,^10,11^ Tanzania,^12^ and Uganda.^13^ Inter-epidemic RVF, while underappreciated relative to regional epidemics, is of significant public health concern given human morbidities imposed by RVFV infection during inter-epidemic periods.^9,11^ Inter- epidemic RVF may also demand a modified disease prediction paradigm: numerous factors besides rainfall, including elevation, topography, soils, and various aspects of human or livestock host populations, plausibly influence inter-epidemic RVF dynamics at fine spatial scales.^5,10,14–16^

Our predictive understanding of RVF is further complicated by global climate change, which promises to shift the risk landscape of numerous infectious diseases.^17^ Critically, climate change can have multifaceted effects on the arthropod vectors of zoonotic diseases, including influences on pathogen transmission efficiency and vector distribution.^18^ As such, climate change is expected to alter RVF’s spatial extent and magnitude of impact.^4,9,19^ However, existing methods for RVF forecasting are primarily focused on near-term prediction of regional epidemics (i.e., they focus on early warning weeks or months ahead of large RVF outbreaks).

These approaches do not allow for RVF risk prediction over the longer timescales relevant to climate change nor do they address disease risk during inter-epidemic periods.^2^ Consequently, we do not yet understand how inter-epidemic RVF risk is likely to change under future climate scenarios, and this knowledge gap hinders effective prioritization of research, policy, and public health practices (e.g., vaccine allocation) that could ensure healthier futures.

Here, we use inter-epidemic RVF outbreak data and machine learning to project inter-epidemic RVF risk under future climate scenarios across Kenya, Tanzania, and Uganda (hereafter, “the study region”). Our specific objectives were to: 1) model historical inter-epidemic RVF outbreaks to learn relationships between environmental features and inter-epidemic RVF risk, 2) validate the inter-epidemic RVF risk model using independent serological data from humans, 3) project inter-epidemic RVF risk under future climate scenarios using learned relationships from the validated model, and 4) estimate the human population at risk from inter-epidemic RVF under future climate scenarios. Ultimately, our analyses identify key abiotic and biotic factors that are associated with inter-epidemic RVF outbreaks and project how changing environmental conditions in the coming decades will translate to changes in the inter-epidemic RVF risk landscape across East Africa.

## METHODS

Our analysis workflow had four discrete steps. First, we used a machine learning model to identify the environmental features that predict historical inter-epidemic RVF outbreaks. Using the model, we were able to retrodict the relative likelihood of inter-epidemic RVF across the study region for all months from 2008-2021. Second, we validated these retrodictions using serological data on RVFV exposure in humans, data which was never used in model training. Third, we used the validated model to develop multi-model projections of inter-epidemic RVF risk across the study region for three future time periods (2021-2040, 2041-2060, and 2061- 2080) under three climate scenarios (SSP126, SSP245, and SSP370; see Supplementary Methods, “The Shared Socioeconomic Pathways”). Finally, we combined our projections of inter-epidemic RVF risk with projections of human population to estimate the population at risk from inter-epidemic RVF under future climate scenarios. We describe each step in greater detail below.

### Modeling inter-epidemic RVF outbreaks

To train a predictive model of inter-epidemic RVF, we required spatially- and temporally-explicit disease presence and absence points, as well as associated predictor variables. Below we describe our procedures for defining RVF presence and absence locations, collating predictor variables, and training the machine learning model.

#### Inter-epidemic RVF outbreak data and background data

To model inter-epidemic RVF risk, we required disease presence and absence data (i.e., outbreak and background points) from across the study region. For disease presence data, we leveraged an existing dataset of 100 human and livestock RVF outbreaks occurring in Kenya, Tanzania, and Uganda from 2008-2022 (Figure 1).^9^ This dataset is composed of small RVF outbreaks (1-129 cases) that occurred during inter-epidemic periods. As such, our work investigates the factors that drive RVF cases, but we do not address the large, multi-country epidemics that often dominate discussion in the RVF literature. For disease absence data, we generated a total of 27,000 background points from across the study region, 150 from each month from 2008-2022 (Figure S1). Critically, to reduce the effects of reporting bias, we followed the approach of Gibb et al.^20^ and generated background points in proportion to human population density (see Supplementary Methods, “Background Data”).

**Figure 1.**
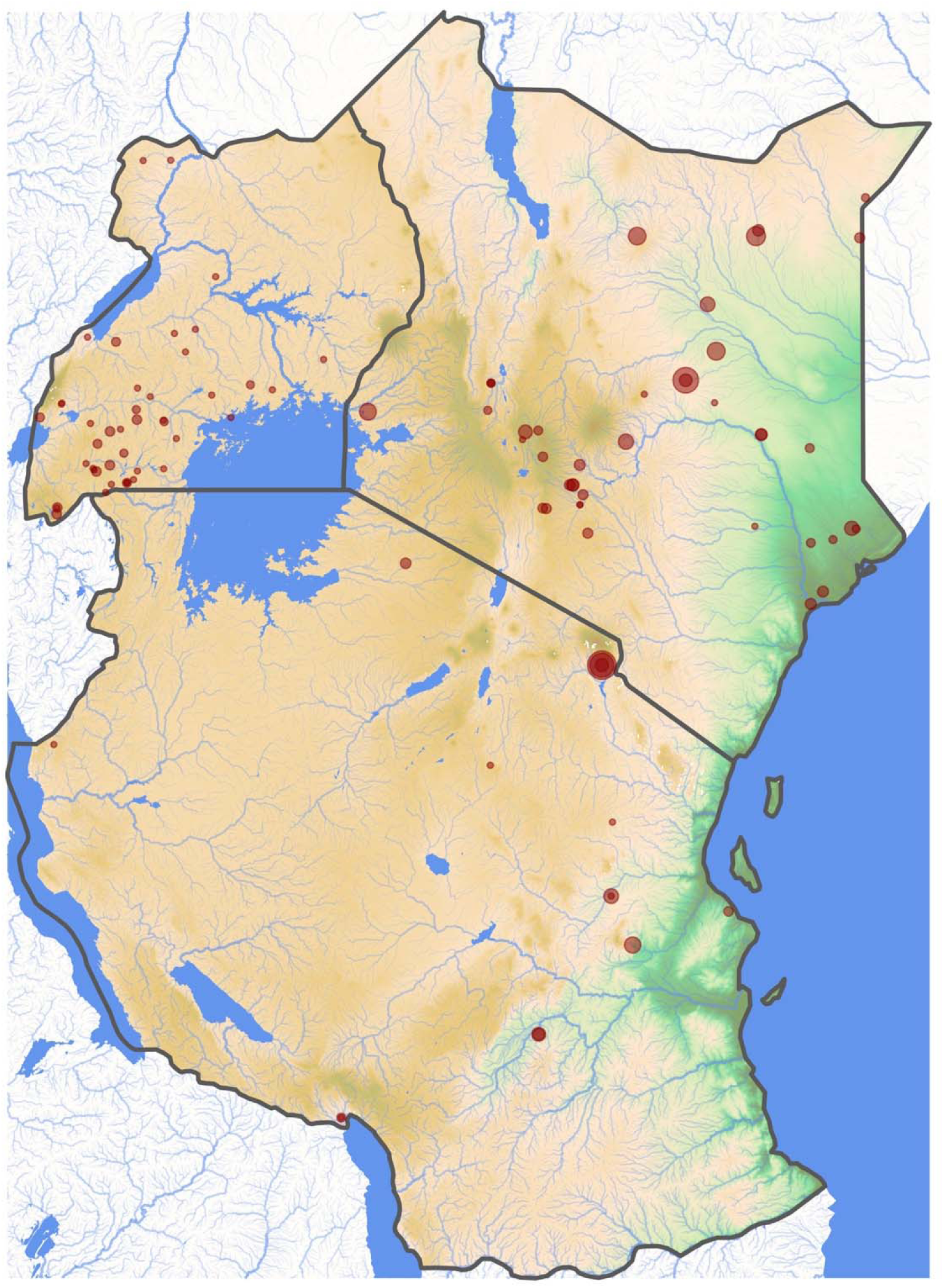
Topographic map of Kenya, Tanzania, and Uganda showing the locations (red points) of 100 inter-epidemic Rift Valley fever outbreak events from 2008-2022. Point size is proportional to the outbreak case count (range: 1-129). Background color represents elevation across the study region, with low elevation in dark green and high elevation in dark brown.

#### Predictor variables

To train our model, we used a suite of 34 spatially-explicit predictors, including variables capturing physical landscape features (e.g., hydrology, soil characteristics, topography), weather (e.g., precipitation, temperature), disease detection processes (e.g., travel time to healthcare), and the abundance of key RVF hosts (e.g., human and livestock density) (Table S1). These predictors have been implicated in driving RVF risk across spatial scales.^5,14,15^ We processed all predictor variable raster layers to a resolution of 2.5 arcminutes (4.63 km x 4.63 km grid cells at the equator). Given this resolution, our study region was covered by a total of 124,313 grid cells. More detail on predictor variables can be found in the Supplementary Methods, “Predictor Variables” section.

#### Model training and evaluation

Once inter-epidemic RVF outbreak points, background points, and predictor variables were collated, we fit gradient-boosted decision trees using XGBoost.^21^ Prior to model training, we split our collated dataset of outbreak and background points into training and test datasets. The training data consisted of all points from 2008-2018 (70 RVF outbreaks/19,800 background). The test dataset consisted of all points from 2019-2022 (30 RVF outbreaks/7,200 background). We tuned model hyperparameters using cross-validation and evaluated model performance on the test dataset using the area under the receiver operating characteristic curve (AUC) measure (see Supplementary Methods, “Model Training”).

Using the trained model, we generated historical predictions (retrodictions) of the relative likelihood of inter-epidemic RVF outbreak across the study region for all months from 2008- 2021. While the model training process only used predictor data from grid cells containing outbreak or background points, retrodictions used predictor data from across the entire study region (i.e., we made retrodictions for every grid cell). Because weather predictor data were unavailable for 2022 (see Supplementary Methods, “Predictor Variables”), we did not make retrodictions for that year. In addition, because we assume the travel time to healthcare predictor only influences disease reporting and not underlying RVF dynamics, we set this value to 0 when generating predictions. We argue this approach better recovers the true RVF risk landscape, with reporting bias factored out. Generating model-based retrodictions from 2008-2021 allowed us to investigate historical temporal and spatial patterns in the relative likelihood of inter-epidemic RVF. We take relative likelihood values (the raw predictive output from our model) as a proxy for inter-epidemic RVF risk and therefore use these terms interchangeably. We performed all model fitting and analysis in R version 4.4.1^22^ using the *tidymodels* package collection.^23^

### Independent model validation

After generating retrodictions, we validated our model’s predictive ability using human serological data, a fully independent data source. We compiled published and unpublished RVFV serological data from humans across our study region,^8,24^ targeting individual-level data where serostatus, location, and age was known. Our data compilation efforts recovered a total of 5,819 serological assays across Kenya (n = 5,277) and Tanzania (n = 542), representing 128 seropositive individuals (2.2% seroprevalence). We assigned each individual to their corresponding 2.5 arcminute grid cell, yielding a total of 406 grid cells with serological data. We then excluded all cells with less than 20 individuals and used the age-structured seroprevalence data from the remaining 103 grid cells (n = 3,031 serological assays) to estimate the local force of infection (FOI).^25^ Having obtained grid cell-level estimates of RVFV FOI, we then calculated the mean grid cell-level relative likelihood of inter-epidemic RVF from 2008-2021 using model-based retrodictions. We evaluated the significance of the relationship between estimated RVFV FOI and model-predicted inter-epidemic RVF likelihood using a linear model with FOI as the outcome, RVF likelihood as the predictor, and the number of serological assays conducted within each grid cell as a weight.

### Projecting inter-epidemic RVF risk under future climate scenarios

To project future inter-epidemic RVF risk, we used the same set of 34 predictors previously described (Table S1), but, instead of historical weather data, we used statistically downscaled, 2.5 arcminute climate projections for three future time periods (2021-2040, 2041-2060, 2061- 2080) under three climate scenarios (SSP126, SSP245, SSP370) (see Supplementary Methods, “Predictor Variables”). For each future scenario, we developed multi-model risk projections across the study region for each month of the calendar year using the outputs of 11 climate models from the Coupled Model Intercomparison Project (CMIP6). In making risk projections, static predictor variables were held constant, human population density varied according to the time period and SSP, and temperature and precipitation predictors varied according to the time period, SSP, climate model, and month. For comparison, we also retrodicted inter-epidemic RVF risk assuming historical climate (1970-2000). Here, we used human population density predictor data from the year 2000, the earliest year available.

### Estimating human population at risk of inter-epidemic RVF under future climate scenarios

Using our projected inter-epidemic RVF risk landscapes, we estimated the total human population at risk of inter-epidemic RVF under future climate scenarios. This involved combining our risk projections with spatially-explicit estimates of future human population density (see Supplementary Methods, “Predictor Variables”). To define areas at risk for inter-epidemic RVF under a given future scenario, we thresholded all 12 monthly risk layers using the value that maximized the true skill statistic (TSS). These thresholded monthly risk layers delineated areas suitable for inter-epidemic RVF. We considered the human population in a given grid cell at risk of inter-epidemic RVF only if the cell was suitable for inter-epidemic RVF for the majority of the calendar year. Using this method, we estimated the human population at risk of inter-epidemic RVF for all future scenarios described above. For comparison, we also estimated the human population at risk assuming historical climate (1970-2000) and human population density from the year 2000.

## RESULTS

### Modeling inter-epidemic RVF outbreaks

Model training produced an XGBoost model with a mean AUC score of 0.84 (SE = 0.06) across the three training data folds. On withheld test data, the model had an AUC of 0.76 (Figure S2), indicating good ability to distinguish RVF outbreak locations from background points. When model predictions were thresholded to maximize the TSS, the model had a TSS of 0.50, with a true negative rate (specificity) of 0.77 and a true positive rate (sensitivity) of 0.73 (Figure S2).

Variable importance for the 34 predictor variables in the model ranged from 0.01 to 0.09 (Figure S3). The five most important predictors were goat density, monthly precipitation, soil silt, elevation, and distance to rivers. Partial dependence plots (PDPs) showed that the relative likelihood of RVF generally decreases with elevation but increases with goat density and soil silt (Figure S4).

Two precipitation variables appeared in the top 10 most important predictors: monthly precipitation in the month of the event and monthly precipitation two months prior (Figure S3). Interestingly, PDPs revealed that precipitation in the month of the event was negatively related to inter-epidemic RVF risk but that the lagged precipitation variable was positively related to risk (Figure S4). While it was less important for prediction, cumulative precipitation in the three months prior to the event was also positively related to RVF (Figure S4). Collectively, these results suggest that RVF risk is elevated during months that have relatively little precipitation but that are preceded by heavy precipitation (i.e., months near the end of rainy seasons).

PDPs also indicated temperature dependencies for RVF. Monthly mean minimum temperatures > 20°C tended to increase disease risk, while monthly mean maximum temperatures > 30°C decreased risk (Figure S4). These results suggest an optimal thermal range for inter-epidemic RVF between ∼20-30°C.

Model-based retrodictions allowed us to examine temporal and spatial patterns in historical inter-epidemic RVF risk across the study region. Despite year-to-year variation, retrodictions showed clear intra-annual seasonal patterns, with the relative likelihood of RVF tending to peak May-July at the end of the long rains (March-May) (Figure 2). Likelihood of inter-epidemic RVF was predicted to be consistently low August-November but was more variable January-April following the cessation of the short rains (October-December) (Figure 2).

**Figure 2.**
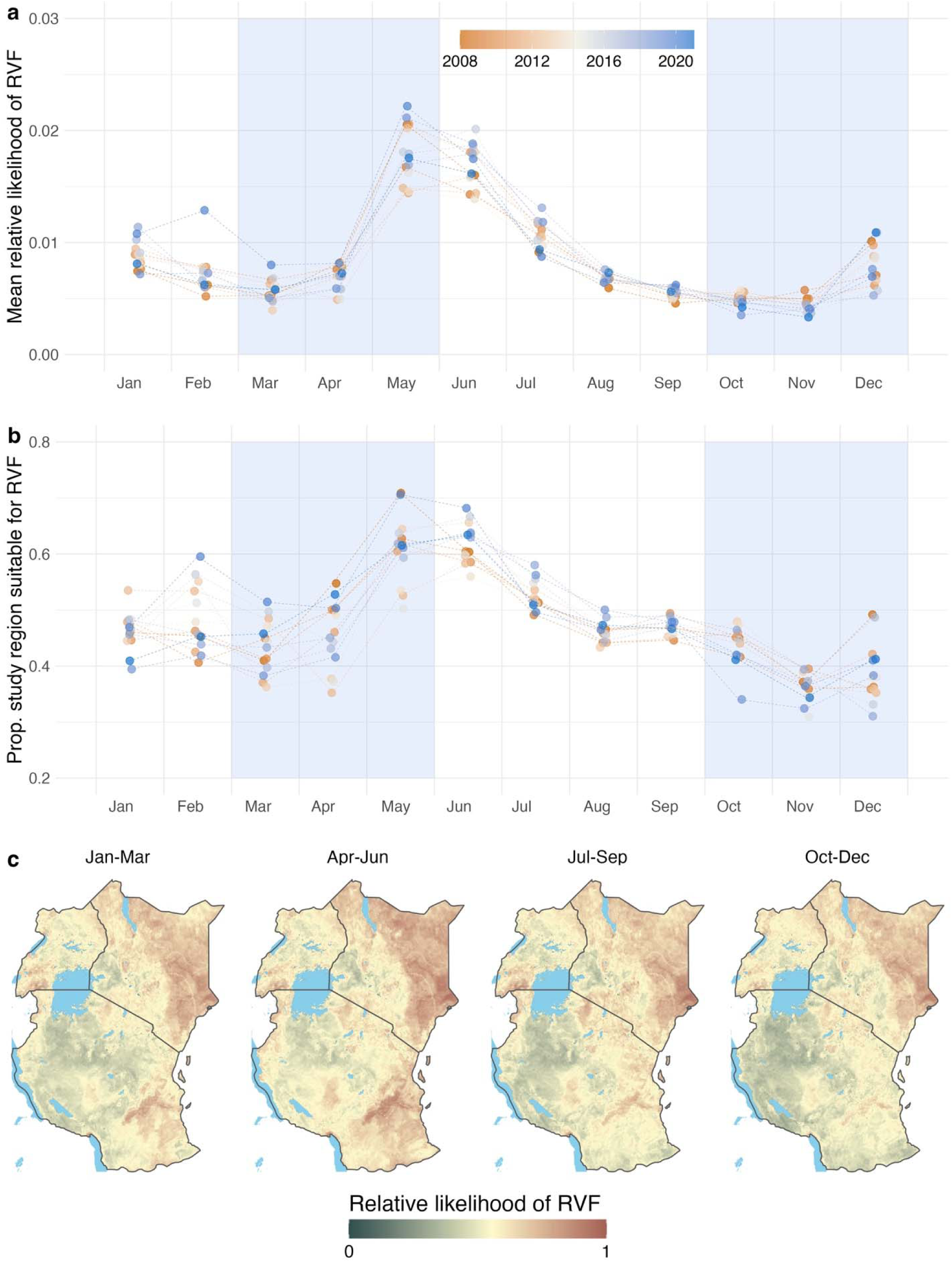
Temporal and spatial patterns in predicted historical inter-epidemic Rift Valley fever risk. Using the fit XGBoost model, we retrodicted inter-epidemic RVF risk for every month from 2008-2021. We summarized model predictions into either the mean relative likelihood of inter-epidemic RVF across the study region (**a**) or the proportion of the study region suitable for RVF when model predictions were thresholded to maximize the true skill statistic (**b**). The colorbar in (**a**) and (**b**) indicates year, with orange representing earlier years and blue representing more recent years. Blue shaded regions in (**a**) and (**b**) represent the long rains (March-May) and short rains (October-December) seasons. We also grouped retrodictions by months and averaged across all years from 2008-2021 to produce seasonal risk maps (**c**). In these maps, we plotted predictions on a log10 scale to better emphasize areas of intermediate risk. In the colorbar in (**c**), dark green corresponds to low relative likelihood of inter-epidemic RVF while dark red corresponds to high likelihood. The colorbar was constructed such that yellow corresponds to the model’s threshold value. In other words, yellow areas represent regions that are on the threshold of suitability for inter-epidemic RVF.

Inter-epidemic RVF risk was concentrated in the low-elevation regions of east Kenya and southeast Tanzania, particularly during the long rains (Figures 2 & S5). There were also notable hotspots of RVF risk in southwest Uganda and the Mbeya Region (Kyela District) of Tanzania north of Lake Malawi (Figure S5). Lower risk of inter-epidemic RVF was predicted for the high-elevation inland regions of Kenya bordering Lake Victoria as well as large swaths of west Tanzania, centered on the Tabora, Shinyanga, and Geita Regions (Figure S5). During the short rains (October-December), RVF risk was noticeably reduced across much of Tanzania while it remained elevated in east Kenya (Figure 2).

### Independent model validation

RVFV force of infection estimated from serosurveys was positively correlated with retrodictions of inter-epidemic RVF likelihood generated using our XGBoost model (β = 0.16, SE = 0.05, p = 0.001; Figure 3). This suggests that our model, which was trained solely on RVF outbreak location data, captures important processes driving human exposure to RVFV.

**Figure 3.**
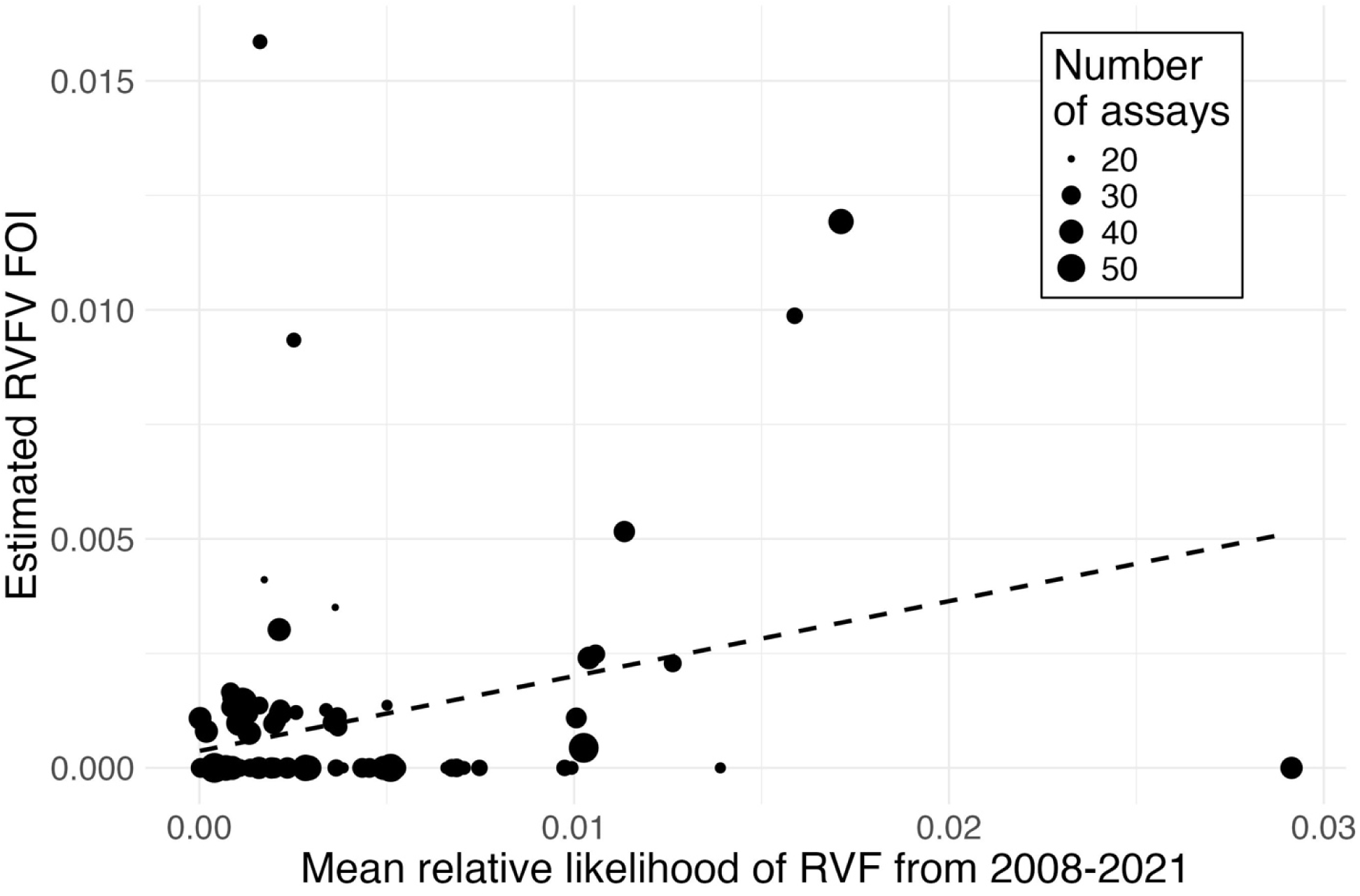
Validation of machine learning predictions of inter-epidemic Rift Valley fever risk. To independently validate the ability of the XGBoost model to predict inter-epidemic RVF risk, we used age-specific serological data from humans to estimate the RVFV force of infection (FOI) across 103 grid cells in our study region. We compared these grid cell-level FOI estimates to the mean model-predicted RVF relative likelihood from 2008-2021 for the same grid cells. Point size is proportional to the number of serological assays conducted within each grid cell, and the dashed line shows the best fit line, weighted by the number of serological assays.

### Projecting inter-epidemic RVF risk under future climate scenarios

Projections of inter-epidemic RVF risk under projected mid- and late-21^st^ century climate conditions also showed seasonality (Figure 4), but with subtle changes relative to historical climate (Figure S5). Most notably, there was increased RVF risk early in the calendar year (January-March), particularly in later time periods (i.e., 2061-2080) and under the high-emissions SSP370 scenario (Figure 4). This trend was largely attributable to expected risk increases in portions of east Kenya. Specifically, the areas roughly concordant with Marsabit, Isiolo, Tana River, and northwestern Garissa Counties were predicted to have increased likelihood of inter-epidemic RVF January-March (Figures S6-8).

**Figure 4.**
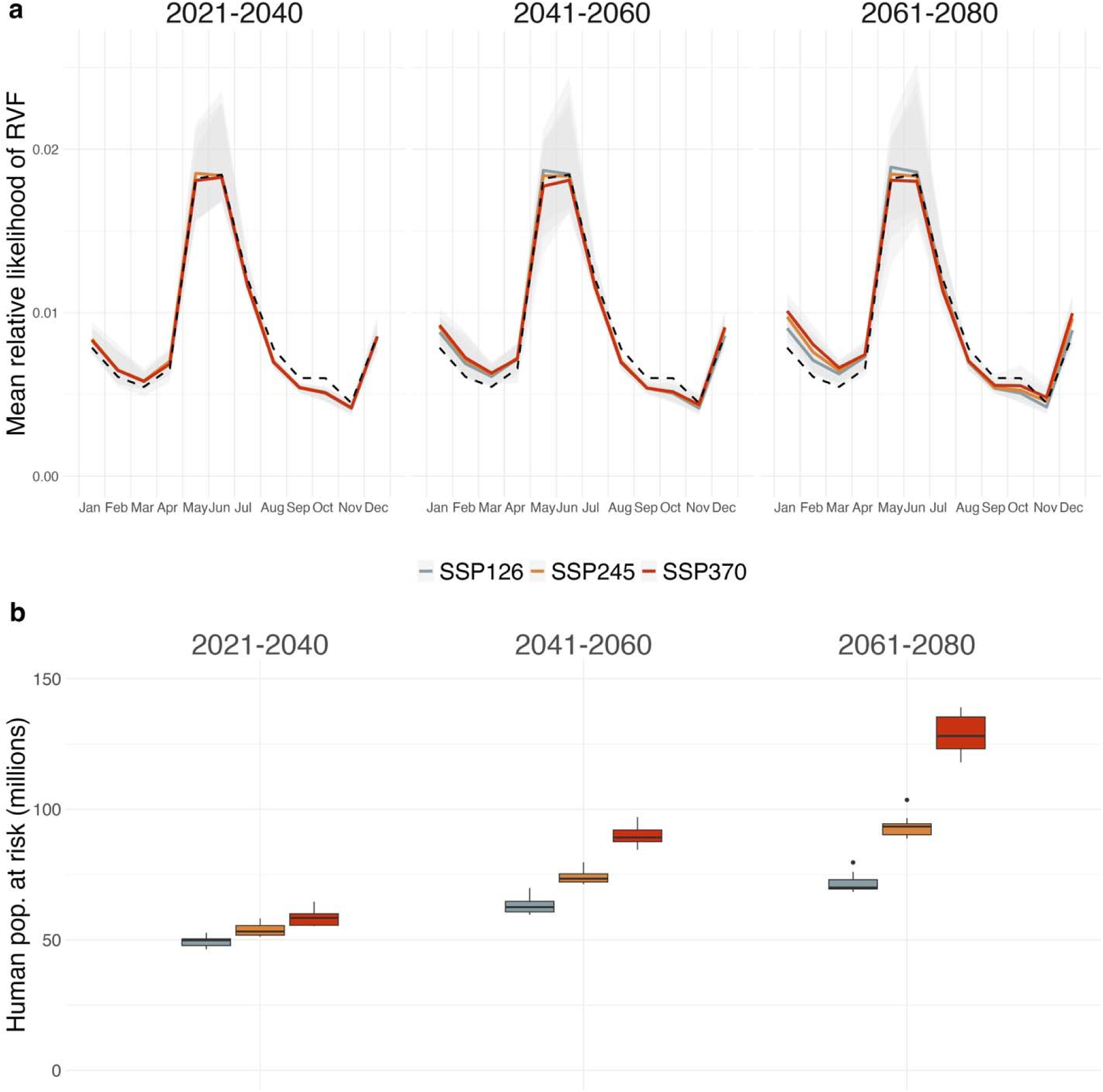
Projected future inter-epidemic Rift Valley fever risk and corresponding human population at risk. Using the fit XGBoost model, we predicted RVF risk under both historical (1970-2000) and future climatic conditions (**a**). The monthly predictions for the single historical climate scenario are shown as a dashed black line in each panel facet to provide a consistent basis for comparison. Projections under future conditions were generated for three time periods (panel columns), three climate scenarios (line color), and 11 climate models. The solid lines within each panel facet correspond to the multi-model mean across the 11 climate models, while the gray shaded regions indicate the range of risk projections across climate models. Multi-model projection ensembles were combined with projections of human population density to estimate the human population exposed to inter-epidemic RVF for the majority of the calendar year under each future scenario (**b**).

Although mean RVF risk in the key high-risk period of May-July appeared relatively stable between historical and future climate scenarios (Figure 4), the mean disguised opposing trends in different parts of the study region. For example, increased RVF risk May-July in southeast Tanzania, specifically the Morogoro Region, was offset by decreased risk relative to historical conditions in the east Kenyan Counties of Tana River, Garissa, and Wajir (Figures S6- 8). In Uganda, the future risk landscape for inter-epidemic RVF is expected to be similar to historical conditions, except for decreases in risk in southwest Uganda in some months (Figures S6-8).

### Estimating human population at risk of inter-epidemic RVF under future climate scenarios

We estimated that under historical climate conditions, ∼25 million people throughout the study region lived in areas with high inter-epidemic RVF risk for most of the calendar year (Table S2). Irrespective of climate scenario, projected increases in human population in the study region translate to increasingly large numbers of people exposed to inter-epidemic RVF under future conditions. By 2061-2080, we estimated that ∼68-139 million people throughout the study region will be living in areas with high inter-epidemic RVF risk throughout most of the year (Figure 4; Table S2). However, population exposure to RVF was highest under SSP370: all estimates for 2061-2080 and this climate scenario indicated > 117 million people at risk, a fourfold increase relative to the historical estimate.

## DISCUSSION

We developed and validated a machine learning model of inter-epidemic RVF that predicted seasonal and spatial variation in disease risk across Kenya, Tanzania, and Uganda. When used to project future RVF risk, the model suggested that risk January-March will increase relative to historical climate, including in the lowest emissions scenarios. Significantly, even during parts of the year when mean disease risk across the study region is predicted to be relatively stable, we recover a shifting risk landscape, with certain areas becoming more suitable for inter-epidemic RVF as others become less suitable. Further, given expected human population growth, we estimate that > 100 million people in the study region may be exposed to inter-epidemic RVF for the majority of each calendar year by 2061-2080. Our results underscore the threat that RVF could pose to public health across East Africa in the coming decades.

Our model indicated that precipitation, soil characteristics, elevation, and goat density were among the most important predictors of inter-epidemic RVF. We found that inter-epidemic RVF outbreaks were more likely to occur in relatively dry months that were preceded by wet months. As such, there was distinct seasonality in predicted inter-epidemic RVF risk, with elevated risk May-July following the long rains (March-May) and a secondary peak December-January following the short rains (October-December). These findings agree with prior work on the timing of RVF outbreaks^16^ and are unsurprising given the key role that precipitation plays in the growth of RVFV vector populations.^3,14,15^ Previous studies also found, as we did, that certain soil types and lower elevations are associated with RVF.^14,15,26^ These environments likely represent optimal habitat for RVFV vectors and cryptic RVFV circulation. Interestingly, we found that goat density was the single most important predictor of inter-epidemic RVF, with higher densities driving increased disease risk. This relationship deserves further investigation, but our work suggests that goats could serve as important RVF sentinels.^27^

Our model also predicted substantial spatial variation in inter-epidemic RVF risk across the study region, and this heterogeneous risk landscape is expected to persist under future climatic conditions. Large portions of low-elevation east Kenya were predicted to be at high risk of inter-epidemic RVF, including the Counties of Marsabit, Isiolo, Tana River, Lamu, Garissa, Wajir, and Mandera. These findings are congruent with prior work demonstrating elevated RVF risk in many of these same areas.^11,14,15^ Crucially, because of our background data selection methods, our model was capable of identifying these sparsely populated Counties as high-risk despite presumably limited disease reporting. Similarly, our dataset contained no RVF events from Turkana County in northwest Kenya, yet our model predicted high suitability for inter-epidemic RVF in this high-risk region.^10,11^ In Tanzania, inter-epidemic RVF risk was primarily concentrated in low-elevation coastal and inland regions of the southeast, namely the Tanga, Pwani, Dar es Salaam, Lindi, and Morogoro Regions as well as the islands of Pemba and Zanzibar. We also recovered a major RVF hotspot immediately north of Lake Malawi in the Mbeya Region (Kyela District), mirroring prior climate-driven modeling.^19^ Human RVFV seroprevalence can approach 30% in the low-lying areas adjacent to Lake Malawi,^12^ and this region of Tanzania deserves special attention as a site of endemic RVFV circulation.^26,28^ Finally, in Uganda, our model suggested high risk in the country’s southwest, an area that is likely an epicenter for inter-epidemic RVFV transmission.^13,29^ Collectively, our results highlight regions to prioritize for RVF surveillance and mitigation both now and in the future.

Projections of inter-epidemic RVF risk showed risk increases January-March in response to anticipated climatic changes. This risk scaled with warming, being most pronounced in the high-emissions SSP370 scenario toward the end of the 21^st^ century. Spatially, this trend manifested most clearly in the Kenyan Counties of Marsabit, Isiolo, and Tana River, suggesting that future climatic changes, including increased short rains (October-December) precipitation^9,30^ and increasing temperatures, will create more favorable conditions for RVF in these arid, low-elevation regions. We emphasize that climate change is expected to reshape the seasonality of RVF risk in some locations. For example, while we predict that Tana River County will become more suitable for inter-epidemic RVF January-March, this same area is predicted to become less suitable May-June. Despite these complexities, a core message of our predictive work is that the eastern lowlands of Kenya are currently at high risk of inter-epidemic RVF and will also undergo climate-driven shifts in disease risk. In Tanzania, the primary finding from our climate modeling was an increase in RVF risk in the Morogoro Region in May and June. The Morogoro Region, which we predict is already at high risk of inter-epidemic RVF and where there is complementary evidence of inter-epidemic RVFV circulation,^28^ deserves increased public health attention. Finally, in Uganda, the only noticeable change in future inter-epidemic RVF risk was a reduction in risk in the southwest. Interestingly, projected risk reductions in this area are a continuation of changes to the risk landscape that are already underway: our retrodictions for 2008-2021 show slightly decreased risk in southwest Uganda when compared to predictions made under 1970-2000 climate (Figure S5). We stress that southwest Uganda remains a high-risk area within the broader study region, even accounting for projected risk decreases.

In sum, our inter-epidemic RVF model shows seasonally varying disease risk, with highest risk January-July. Risk is projected to increase January-March under future climate scenarios. These predictions are particularly concerning because, when coupled with projections of human population growth, they suggest that > 100 million people in the study region may be exposed to inter-epidemic RVF by 2061-2080. Consequently, we urge increased disease surveillance, prevention, and control efforts in the geographic areas we identify as high-risk.

Although different climate futures do not imply radically different RVF risk landscapes in our analyses, the important predictive role of goat density suggests that livestock abundance and management practices may serve as key moderators of disease risk. Further work at the animal-human interface would clarify the specific conditions that lead to human RVFV exposure, and targeted public health interventions guided by predictive models could help mitigate RVF’s human and animal health impacts in a changing climate.

## Data Availability

Data and code supporting this manuscript are publicly available via GitHub (https://github.com/eveskew/RVFutures).

## Contributors

EAE: conceptualisation, data curation, formal analysis, methodology, validation, writing – original draft; EC: conceptualisation, data curation, methodology, validation, writing – review & editing; DS: conceptualisation, methodology, writing – review & editing; SS: data curation, methodology, writing – review & editing; LN: methodology, writing – review & editing; MKN: conceptualisation, funding acquisition, methodology, project administration, supervision, writing – review & editing; SLN: conceptualisation, funding acquisition, methodology, project administration, supervision, validation, writing – review & editing. EAE, EC, SS, and SLN have directly accessed and verified the underlying data. All authors gave final approval for publication.

## Acknowledgements

Funding for the project was provided by the US National Institute of Allergy and Infectious Diseases/National Institutes of Health (NIAID/NIH), grant number U01AI151799 through the Center for Research in Emerging Infectious Diseases-East and Central Africa (CREID-ECA) and grant number R01GM122079 to SLN. Serological data and metadata were provided by the University of Glasgow and were generated through the Zoonoses and Emerging Livestock Systems program funded by the UK Biotechnology and Biological Sciences Research Council (BB/L018926/1 and BB/J010367).

