## Supplementary Methods for "Projecting climate change impacts on inter-epidemic risk of Rift Valley fever across East Africa"

Journal: *The Lancet Planetary Health*

Authors:

Evan A. Eskew (corresponding author)

Affiliations: Institute for Interdisciplinary Data Sciences, University of Idaho, Moscow ID, USA

Erin Clancey

Affiliations: Paul G. Allen School for Global Health, Washington State University, Pullman WA, USA

Deepti Singh

Affiliations: School of the Environment, Washington State University, Vancouver WA, USA

Silvia Situma

Affiliations: Washington State University Global Health-Kenya, Nairobi, Kenya; Department of Animal Science, Pwani University, Kilifi, Kenya

Luke Nyakarahuka

Affiliations: Department of Biosecurity, Ecosystems & Veterinary Public Health, Makerere University, Kampala, Uganda; Uganda Virus Research Institute, Entebbe, Uganda

M. Kariuki Njenga

Affiliations: Paul G. Allen School for Global Health, Washington State University, Pullman WA, USA; Washington State University Global Health-Kenya, Nairobi, Kenya

Scott L. Nuismer

Affiliations: Department of Biological Sciences, University of Idaho, Moscow ID, USA

**TABLE OF CONTENTS**

**Part 1: Supplementary Methods……………………….....…………………………………….2**

**Part 2: Supplementary Tables…………………………………………………………………10**

**Part 3: Supplementary Figures………………………….……………………………………..14**

**Part 4: References………..……………………………………………………………………..27**

**PART 1: SUPPLEMENTARY METHODS**

**The Shared Socioeconomic Pathways**

Projecting future inter-epidemic Rift Valley fever (RVF) risk required a well-defined set of assumptions about what the future world will be like. Here, we relied on the shared socioeconomic pathways (SSPs) framework, a set of standardized modeling scenarios developed by the climate change research community.^1–3^ We focused on three SSP scenarios that each assume a different path of global socioeconomic development: SSP1 (“Sustainability”), SSP2 (“Middle of the road”), and SSP3 (“Regional rivalry”).^2^ SSP1 imagines a world that centers environmental sustainability where there are low societal challenges to both climate change mitigation and adaptation. SSP2 assumes a world that largely follows historical trends, with relatively slow progress on sustainable development goals and intermediate challenges to both climate change mitigation and adaptation. SSP3 envisions a world with a continued reliance on fossil fuels, a lack of international cooperation on environmental problems, and generally ineffective civic institutions, creating a situation with high challenges to both climate change mitigation and adaptation. The development futures imagined by the SSPs are partially independent from the greenhouse gas mitigation scenarios described by the complementary representative concentration pathways (RCPs) framework.^1,3^ A single SSP can be consistent with different RCPs, and, conversely, a given RCP can be consistent with multiple SSPs. For specificity, the three future climate scenarios we considered are paired combinations of SSPs and RCPs, all representing Tier 1 scenarios from the Scenario Model Intercomparison Project, namely SSP1-RCP2.6 (SSP126), SSP2-RCP4.5 (SSP245), and SSP3-RCP7.0 (SSP370).^1^ We explicitly avoided analysis of the SSP5-RCP8.5 “worst-case” climate scenario, as this is increasingly regarded as a very unlikely climate future.^4^ Rather, in our analysis, SSP370 represents the most extreme scenario where the absence of meaningful climate mitigation policy leads to substantial warming.

**Background Data**

The selection of background (i.e., “pseudo-absence”) data for use in machine learning models is a key step with a major impact on model validity and interpretability.^5^ Critically, background data must be selected in light of the biases that shape the observed occurrence data.^6^ In disease modeling, a primary concern is the potential influence of disease reporting bias. Because disease-related outcomes such as pathogen detection or clinical disease are only reported in areas where there is the requisite scientific capacity to surveil, diagnose, and communicate disease events, the observed distribution of disease may be in large part a reflection of underlying disease reporting effort. When widespread reporting biases do exist, the use of random background points for comparison with disease-associated locations can be highly misleading.^6^ Because randomly selected background points will include portions of the study area in which there is very little or no disease reporting effort (and therefore no possibility of observed disease), such an analysis can, in effect, end up distinguishing between areas for which there is disease-related reporting effort (i.e., the disease locations) versus those where there is none. A better approach is to generate background data with a sample selection bias that matches the occurrence data.^6^

In light of these issues, we followed the approach of Gibb et al.^7^ and generated background points in proportion to human population density across the study region. In other words, our background data were preferentially drawn from areas with higher human population density, working under the assumption that these areas have higher disease reporting effort. As a result, our background points represent locations where disease reporting should have been possible, yet no disease reporting occurred. More specifically, for each month from 2008-2022 we selected 150 background points in proportion to the human population density across the study region (Figure S1). This approach ensured that our model’s training data was sufficiently broad with respect to environmental features across space and time (i.e., the model training data included background points from months and years when RVF outbreaks were not observed). We used the WorldPop dataset as our measure of historical human population density across the study region.^8^ Finally, within years, we ensured that background points never occurred within grid cells where inter-epidemic RVF outbreaks were also observed (i.e., a grid cell could never be categorized as both an RVF outbreak and background location in the same year). In total, we generated 27,000 population-weighted background points (1,800 each year from 2008-2022) for use in machine learning modeling (Figure S1). Because of our approach to background data selection, our modeling distinguishes areas where inter-epidemic RVF outbreaks were reported from areas where inter-epidemic RVF outbreaks were not reported but presumably would have been, had they occurred. As such, we expect to more effectively recover the true underlying environmental and biological drivers of inter-epidemic RVF outbreaks having mitigated the effects of reporting bias. If the bias in background data selection appropriately mimics the bias in the observed outbreak data, we do not expect to see predictor variables that should be strong proxies for disease reporting effort (e.g., travel time to healthcare, human population density) play an important role in prediction.

**Predictor Variables**

We used 34 spatially-explicit predictor variables in machine learning modeling of inter-epidemic RVF risk (Table S1). There were three major categories of predictor variables: those that were static over time, those that varied yearly, and those that varied monthly. Here, we describe the predictor variables in detail. For predictors that varied over time, we include an explanation of data sourcing for both historical and future time periods.

*Static Predictor Variables*

Twenty of our predictor variables were static over time (Table S1). These included five hydrology variables, nine soil variables, two topographic variables, travel time to healthcare, and three livestock density variables. For hydrology data, we relied on the HydroLAKES^9^ and HydroRIVERS^10^ datasets provided by the HydroSHEDS project (https://www.hydrosheds.org/). Because RVF cases might be more likely to occur near the water features that provide mosquito breeding habitat^11,12^ and given evidence from our study region showing increased human RVFV seroprevalence in areas close to water,^13^ we were interested in calculating distances to hydrological features. Specifically, we used the HydroLAKES data to generate raster layers representing the distance from each grid cell centroid to lakes of four different surface area size classes: lakes of any size, lakes ≥ 1 km^2^, lakes ≥ 5 km^2^, and lakes ≥ 10 km^2^. Using the HydroRIVERS data, we calculated the fifth hydrological variable: the distance from each grid cell centroid to the closest river with a flow rate ≥ 10 m^3^/s. We hypothesized that these larger rivers would be associated with floodplain habitat that could support mosquito populations. To characterize soils across the study region, we downloaded nine variables from the 250 m resolution SoilGrids dataset^14^: bulk density, cation exchange capacity, volumetric fraction of coarse fragments, proportion of clay particles, total nitrogen, soil pH, proportion of sand particles, proportion of silt particles, and soil organic carbon content. Our two topographic variables were elevation and slope. For elevation data, we used the Shuttle Radar Topography Mission (SRTM) dataset,^15,16^ which is natively provided at 1 arcsecond resolution. Slope was then calculated from this elevation data using the ‘terrain()’ function in the R package *terra*.^17^ Travel time to healthcare data came from a dataset quantifying access time to healthcare, assuming the availability of motorized transport.^18^ Finally, for livestock density data, we used the most recent version (version 4) of the Gridded Livestock of the World (GLW) database.^19^ These data were estimates of the global distribution of cattle, goats, and sheep in the year 2015. Data were provided at 5 arcminute resolution. We used the GLWv4 areal-weighted (AW) estimates, and converted livestock count estimates to livestock densities prior to modeling.

*Yearly Predictor Variables*

Human population density was our only predictor variable that varied by year (Table S1). We used human population density data from the WorldPop (https://hub.worldpop.org) dataset,^8^ which provides density estimates for all countries in the study region at 30 arcsecond resolution from 2000-2020. For human population density values for 2021 and 2022, where WorldPop data was not available, we used the estimate for the year 2020. For modeling of future inter-epidemic RVF risk, we used projected human population data for the years 2030, 2050, and 2070, which correspond to the midpoints of the time periods 2021-2040, 2041-2060, and 2061-2080.^20^ These human population projections build on and are validated against the WorldPop dataset, are provided at 30 arcsecond resolution, and include estimates for SSP scenarios 1-5.

*Monthly Predictor Variables*

Thirteen of our predictor variables varied by month (Table S1). For predictive modeling of historical inter-epidemic RVF outbreaks, we relied on historical monthly weather data at 2.5 arcminute resolution provided by WorldClim (<https://www.worldclim.org/data/monthlywth.html>). These data were generated by downscaling the CRU-TS-4.06 dataset^21^ using the WorldClim version 2.1 data for bias correction.^22^ We downloaded monthly precipitation, monthly minimum temperature, and monthly maximum temperature data for the entire historical period of interest (2008-2021; note that WorldClim monthly weather data were unavailable for the year 2022). From these data, we generated a total of five precipitation predictor variables to be associated with each outbreak or background point: monthly precipitation in the month of the event, monthly precipitation one month prior, monthly precipitation two months prior, monthly precipitation three months prior, and cumulative precipitation in the prior three months. Similarly, we generated a total of eight temperature predictor variables: monthly minimum temperature and monthly maximum temperature in the month of the event as well as lagged versions of these two variables at one month, two month, and three month lags (Table S1).

To project inter-epidemic RVF risk under future climate conditions, we required future analogs of the thirteen monthly weather variables described above. Here, we relied on the 2.5 arcminute resolution future climate data provided by WorldClim (<https://www.worldclim.org/data/cmip6/cmip6climate.html>). More specifically, these data were calibrated, downscaled versions of future climate projections derived from climate models included in the Coupled Model Intercomparison Project Phase 6 (CMIP6).^23^ As with the historical monthly weather data described above, these future climate data were downscaled and bias-corrected using WorldClim version 2.1.^22^ From the WorldClim future climate data archive, we downloaded average monthly values of precipitation, minimum temperature, and maximum temperature that varied across three important data dimensions: time period, climate scenario (i.e., SSP scenario), and climate model. First, we downloaded data representing three different 20-year time periods: 2021-2040, 2041-2060, and 2061-2080. Second, we downloaded data for the three SSP scenarios of interest, namely SSP126, SSP245, and SSP370 (see Supplementary Methods, “The Shared Socioeconomic Pathways”). Finally, to account for uncertainty among climate models, we downloaded data from all available CMIP6 climate models on WorldClim that covered the time periods and climate scenarios of interest. As a result, we included climate data from 11 different climate models: ACCESS-CM2, BCC-CSM2-MR, CMCC-ESM2, EC-Earth3-Veg, GISS-E2-1-G, INM-CM5-0, IPSL-CM6A-LR, MIROC6, MPI-ESM1-2-HR, MRI-ESM2-0, and UKESM1-0-LL. In sum, we downloaded monthly precipitation, monthly minimum temperature, and monthly maximum temperature for 99 different combinations of future time period, future climate scenario (i.e., SSP), and climate model (3 time periods x 3 climate scenarios x 11 climate models). Once these three core variables were obtained for each combination, we were able to calculate the full suite of 13 monthly predictors, as described above for the historical weather data, for use in inter-epidemic RVF risk projection.

Finally, we recognized that it would be most relevant to compare projections of inter-epidemic RVF risk under future climate conditions to analagous predictions made using historical climate data. Because the future risk projections are made using future climate data, which necessarily represent long-term averages in weather, they are not directly comparable to our retrodictions of inter-epidemic RVF risk for the 2008-2021 time period, which were generated using the WorldClim historical monthly weather data. Therefore, to generate retrodictions of inter-epidemic RVF risk under historical climate, we used the WorldClim version 2.1 historical climate data (<https://www.worldclim.org/data/worldclim21.html>), which represents the time period 1970-2000.^22^ As with historical weather data and future climate data, for these historical climate data we downloaded monthly precipitation, monthly minimum temperature, and monthly maximum temperature data at the 2.5 arcminute resolution and processed these core variables to derive the full suite of 13 monthly predictor variables.

**Model Training**

As the first step in model training, we tuned XGBoost hyperparameters. We tuned seven hyperparameters: learning rate (range: 0.0001-0.1), minimum number of data points required for node splitting (range: 2-20), number of randomly sampled predictors at each split (range: 2-20), proportion of the total sample used at each model iteration (range: 0.1-0.9), the early stopping parameter (range: 3-20), tree depth (range: 1-3), and number of trees (range: 10-1,000). We specifically implemented early stopping and limited our models to a modest tree depth to control overfitting. We used the ‘grid_max_entropy()’ function from the *dials* R package^24^ to generate 250 parameter combinations that explored the full hyperparameter space. We then conducted hyperparameter tuning by dividing the training dataset into three folds and using cross-validation procedures for these 250 hyperparameter combinations. We considered the hyperparameter combination that led to the highest average area under the receiver operating characteristic curve (AUC) score across the three training data folds to be the best hyperparameter combination. With the best hyperparameter combination identified, we then finalized the model fit on the entirety of the training data and evaluated model performance on the test dataset, again using the AUC measure.

**PART 2: SUPPLEMENTARY TABLES**

**Table S1. Summary of all 34 predictor variables used in XGBoost model training.** For predictors with a yearly or monthly temporal resolution, the “Data Source” column indicates where both historical and future projected data were obtained.

| **Variable Name** | **Variable Type** | **Temporal Resolution** | **Data Source** |
| --- | --- | --- | --- |
| dist. to lake (> 1 km^2) | proximity to water | fixed | HydroLAKES  (<https://www.hydrosheds.org/products/hydrolakes>) |
| dist. to lake (> 5 km^2) | proximity to water | fixed | HydroLAKES  (<https://www.hydrosheds.org/products/hydrolakes>) |
| dist. to lake (> 10 km^2) | proximity to water | fixed | HydroLAKES  (<https://www.hydrosheds.org/products/hydrolakes>) |
| dist. to lake (any size) | proximity to water | fixed | HydroLAKES  (<https://www.hydrosheds.org/products/hydrolakes>) |
| dist. to river (> 10 m^3/s) | proximity to water | fixed | HydroRIVERS  (<https://www.hydrosheds.org/products/hydrorivers>) |
| bulk density | soils | fixed | SoilGrids  (<https://soilgrids.org/>) |
| cation exchange capacity | soils | fixed | SoilGrids  (<https://soilgrids.org/>) |
| volumetric fraction of coarse fragments | soils | fixed | SoilGrids  (<https://soilgrids.org/>) |
| proportion of clay particles | soils | fixed | SoilGrids  (<https://soilgrids.org/>) |
| total nitrogen | soils | fixed | SoilGrids  (<https://soilgrids.org/>) |
| pH | soils | fixed | SoilGrids  (<https://soilgrids.org/>) |
| proportion of sand particles | soils | fixed | SoilGrids  (<https://soilgrids.org/>) |
| proportion of silt particles | soils | fixed | SoilGrids  (<https://soilgrids.org/>) |
| organic carbon content | soils | fixed | SoilGrids  (<https://soilgrids.org/>) |
| elevation | topography | fixed | Shuttle Radar Topography Mission  (<https://www.usgs.gov/centers/eros/science/usgs-eros-archive-digital-elevation-shuttle-radar-topography-mission-srtm-1>) |
| slope | topography | fixed | Calculated in R using elevation data |
| travel time to healthcare | disease detection | fixed | Weiss et al., 2020  (<https://doi.org/10.1038/s41591-020-1059-1>) |
| cattle density | livestock density | fixed | Gilbert et al., 2018  (<https://doi.org/10.1038/sdata.2018.227>) |
| goat density | livestock density | fixed | Gilbert et al., 2018  (<https://doi.org/10.1038/sdata.2018.227>) |
| sheep density | livestock density | fixed | Gilbert et al., 2018  (<https://doi.org/10.1038/sdata.2018.227>) |
| human population density | human density | yearly | 2008-2020:  WorldPop  (<https://hub.worldpop.org>)  2030, 2050, 2070:  Wang et al., 2022  (<https://doi.org/10.1038/s41597-022-01675-x>) |
| monthly precipitation  (month of event) | precipitation | monthly | Historical (2008-2021)  weather data:  WorldClim  (<https://www.worldclim.org/data/monthlywth.html>)  Historical (1970-2000)  climate data:  WorldClim  (<https://www.worldclim.org/data/worldclim21.html>)  Future (2030, 2050, 2070)  climate data:  WorldClim  (<https://www.worldclim.org/data/cmip6/cmip6climate.html>) |
| monthly precipitation  (1 month prior) | precipitation | monthly | Calculated in R using monthly precipitation data |
| monthly precipitation  (2 months prior) | precipitation | monthly | Calculated in R using monthly precipitation data |
| monthly precipitation  (3 months prior) | precipitation | monthly | Calculated in R using monthly precipitation data |
| cumulative precipitation  (3 months prior) | precipitation | monthly | Calculated in R using monthly precipitation data |
| monthly max temperature (month of event) | temperature | monthly | Historical (2008-2021)  weather data:  WorldClim  (<https://www.worldclim.org/data/monthlywth.html>)  Historical (1970-2000)  climate data:  WorldClim  (<https://www.worldclim.org/data/worldclim21.html>)  Future (2030, 2050, 2070)  climate data:  WorldClim  (<https://www.worldclim.org/data/cmip6/cmip6climate.html>) |
| monthly max temperature (1 month prior) | temperature | monthly | Calculated in R using monthly max temperature data |
| monthly max temperature (2 months prior) | temperature | monthly | Calculated in R using monthly max temperature data |
| monthly max temperature (3 months prior) | temperature | monthly | Calculated in R using monthly max temperature data |
| monthly min temperature (month of event) | temperature | monthly | Historical (2008-2021)  weather data:  WorldClim  (<https://www.worldclim.org/data/monthlywth.html>)  Historical (1970-2000)  climate data:  WorldClim  (<https://www.worldclim.org/data/worldclim21.html>)  Future (2030, 2050, 2070)  climate data:  WorldClim  (<https://www.worldclim.org/data/cmip6/cmip6climate.html>) |
| monthly min temperature (1 month prior) | temperature | monthly | Calculated in R using monthly min temperature data |
| monthly min temperature (2 months prior) | temperature | monthly | Calculated in R using monthly min temperature data |
| monthly min temperature (3 months prior) | temperature | monthly | Calculated in R using monthly min temperature data |

**Table S2. Human population in Kenya, Tanzania, and Uganda at risk of inter-epidemic Rift Valley fever under historical and future climates.** The total human population at risk of inter-epidemic RVF was calculated using historical climate data (1970-2000) as well as future climate data for three time periods (2021-2040, 2041-2060, 2061-2080) and three climate scenarios (SSP126, SSP245, SSP 370). For future scenarios, the population at risk and proportion of population at risk columns show both the mean and range of values across 11 climate models.

| **Time Period** | **Climate**  **Scenario** | **Population Size** | **Population at Risk**  **of Inter-epidemic RVF** | **Prop. Population at Risk**  **of Inter-epidemic RVF** |
| --- | --- | --- | --- | --- |
| 1970-2000 | N/A | 86,338,616 | 25,176,220 | 0.29 |
| 2021-2040 | SSP126 | 182,237,790 | 49,419,352  (46,375,252 – 52,704,049) | 0.27  (0.25 – 0.29) |
| 2021-2040 | SSP245 | 195,769,209 | 53,754,679  (51,159,196 – 58,179,199) | 0.27  (0.26 – 0.30) |
| 2021-2040 | SSP370 | 211,999,622 | 58,331,539  (55,218,389 – 64,606,934) | 0.28  (0.26 – 0.30) |
| 2041-2060 | SSP126 | 234,273,097 | 63,063,416  (59,695,563 – 69,858,454) | 0.27  (0.25 – 0.30) |
| 2041-2060 | SSP245 | 276,471,532 | 74,040,186  (71,330,362 – 79,727,286) | 0.27  (0.26 – 0.29) |
| 2041-2060 | SSP370 | 333,370,478 | 89,913,949  (84,501,459 – 97,011,566) | 0.27  (0.25 – 0.29) |
| 2061-2080 | SSP126 | 262,989,553 | 71,718,244  (68,328,998 – 79,644,015) | 0.27  (0.26 – 0.30) |
| 2061-2080 | SSP245 | 341,313,107 | 93,354,172  (88,852,810 – 103,593,471) | 0.27  (0.26 – 0.30) |
| 2061-2080 | SSP370 | 458,941,074 | 129,152,936  (117,969,408 – 139,073,004) | 0.28  (0.26 – 0.30) |

**PART 3: SUPPLEMENTARY FIGURES**

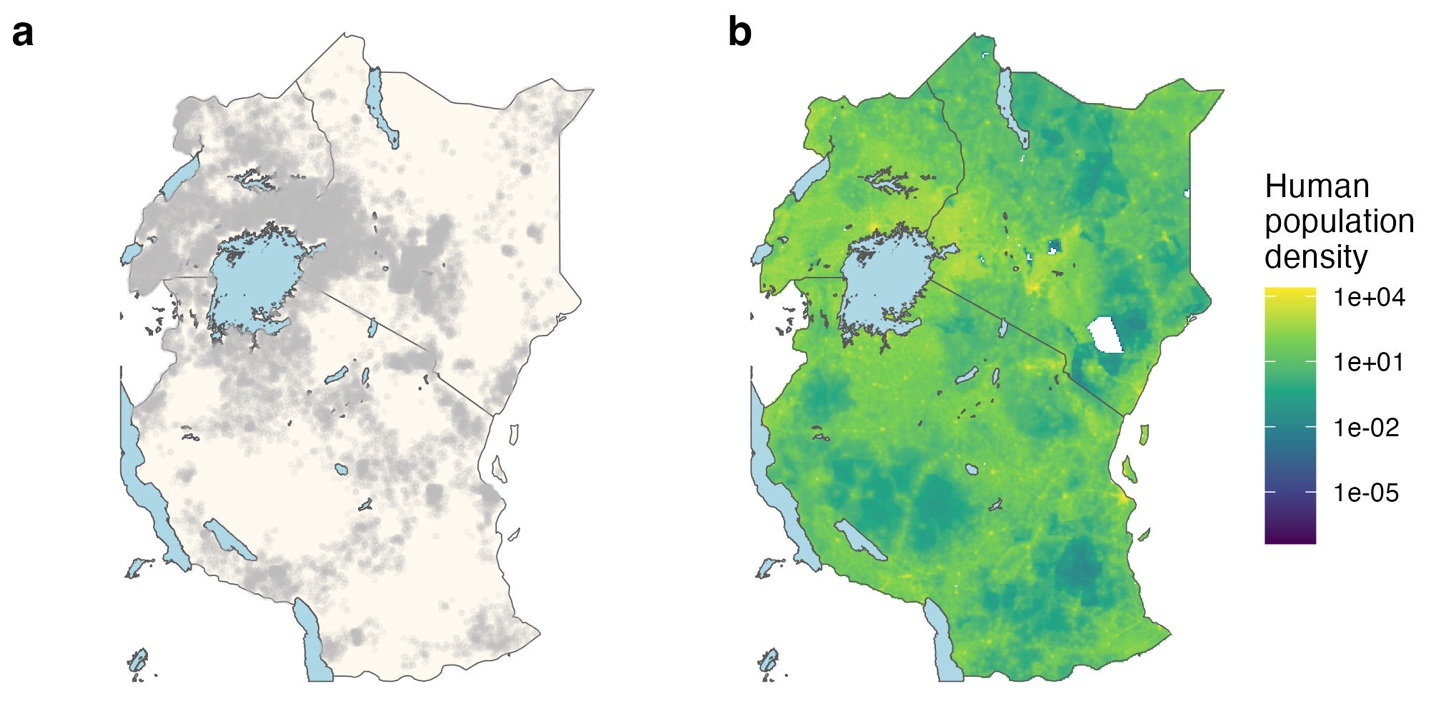

**Figure S1. Background data selection for use in XGBoost modeling.** Panel **a** shows the 27,000 background points (150 for each month from 2008-2022) that were generated in proportion to human population density across the study region. For reference, panel **b** shows mean human population density from 2008-2020 (the available years of the WorldPop human population reference dataset) across the study region. The colorbar in **b** shows human population density data (persons/km^2^) on a log10 scale.

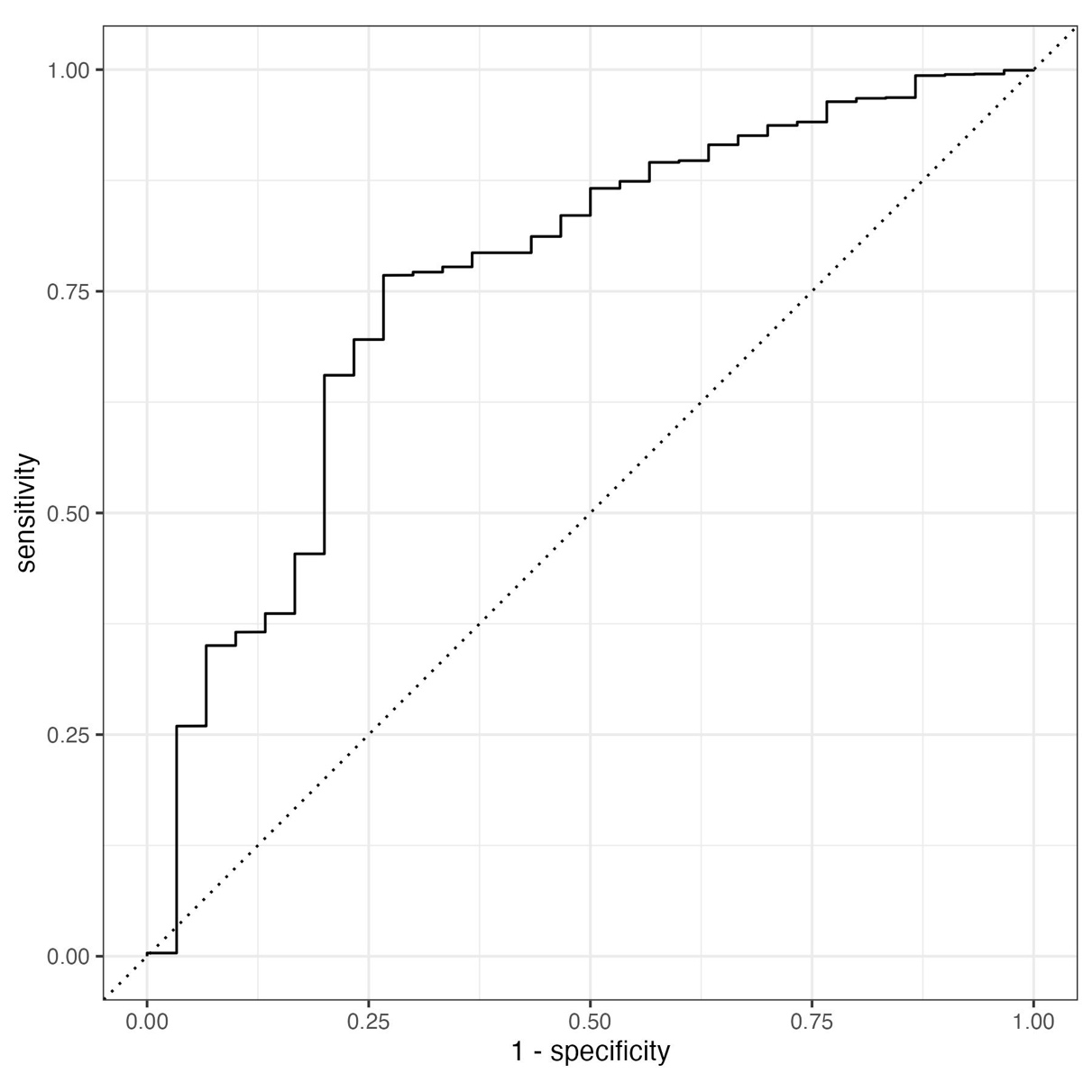

**Figure S2. Receiver operating characteristic curve for the fit XGBoost model on withheld testing data on inter-epidemic RVF outbreaks.** The plot’s y-axis is equivalent to the true positive rate, while the x-axis is equivalent to the false positive rate. The area under the curve (AUC) is 0.76.

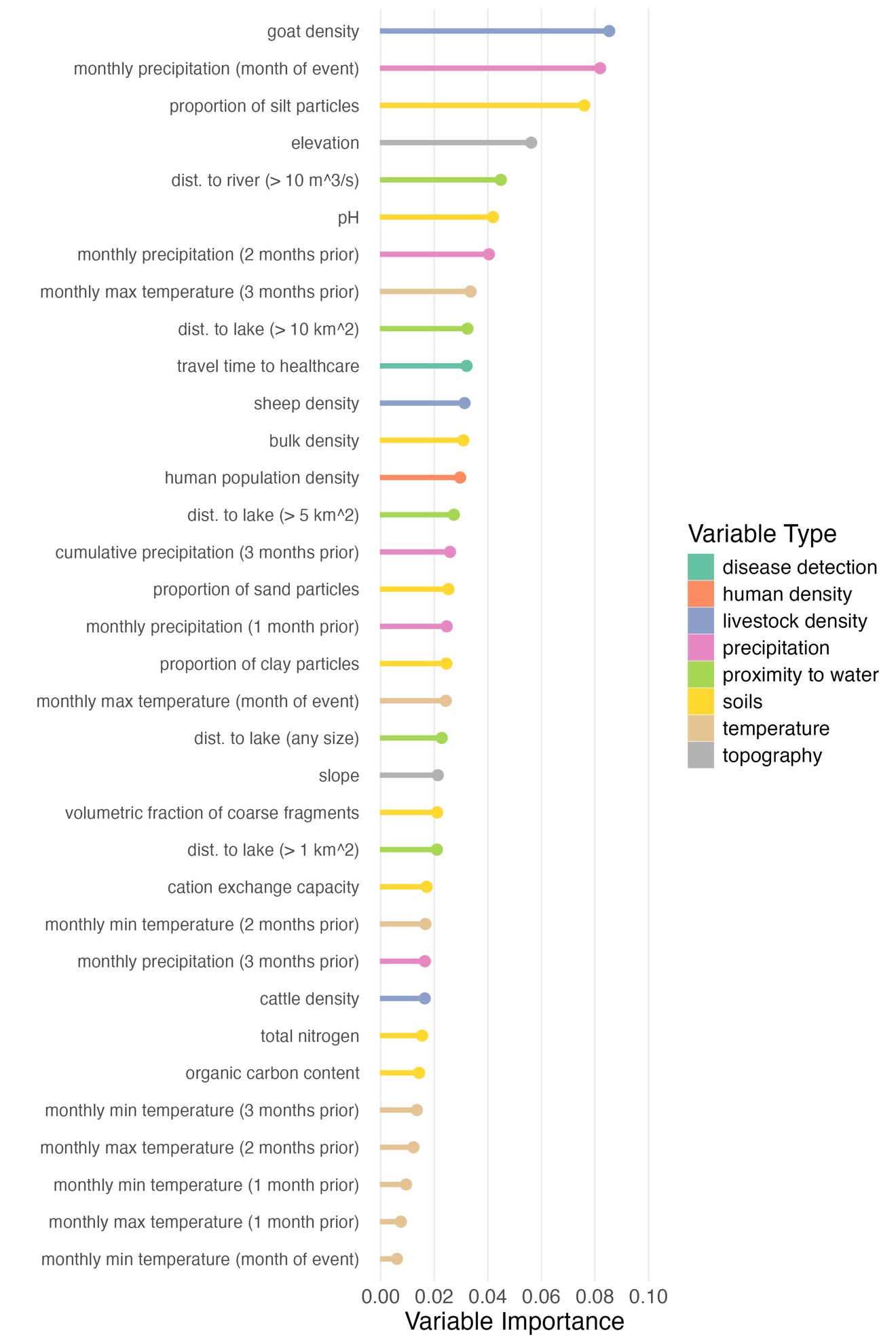

**Figure S3. Variable importance for all 34 predictor variables used in XGBoost model training.** Variables are colored according to their variable type. Note that if all variables were equally important for prediction, the variable importance for all variables would be 1/34 = 0.03.

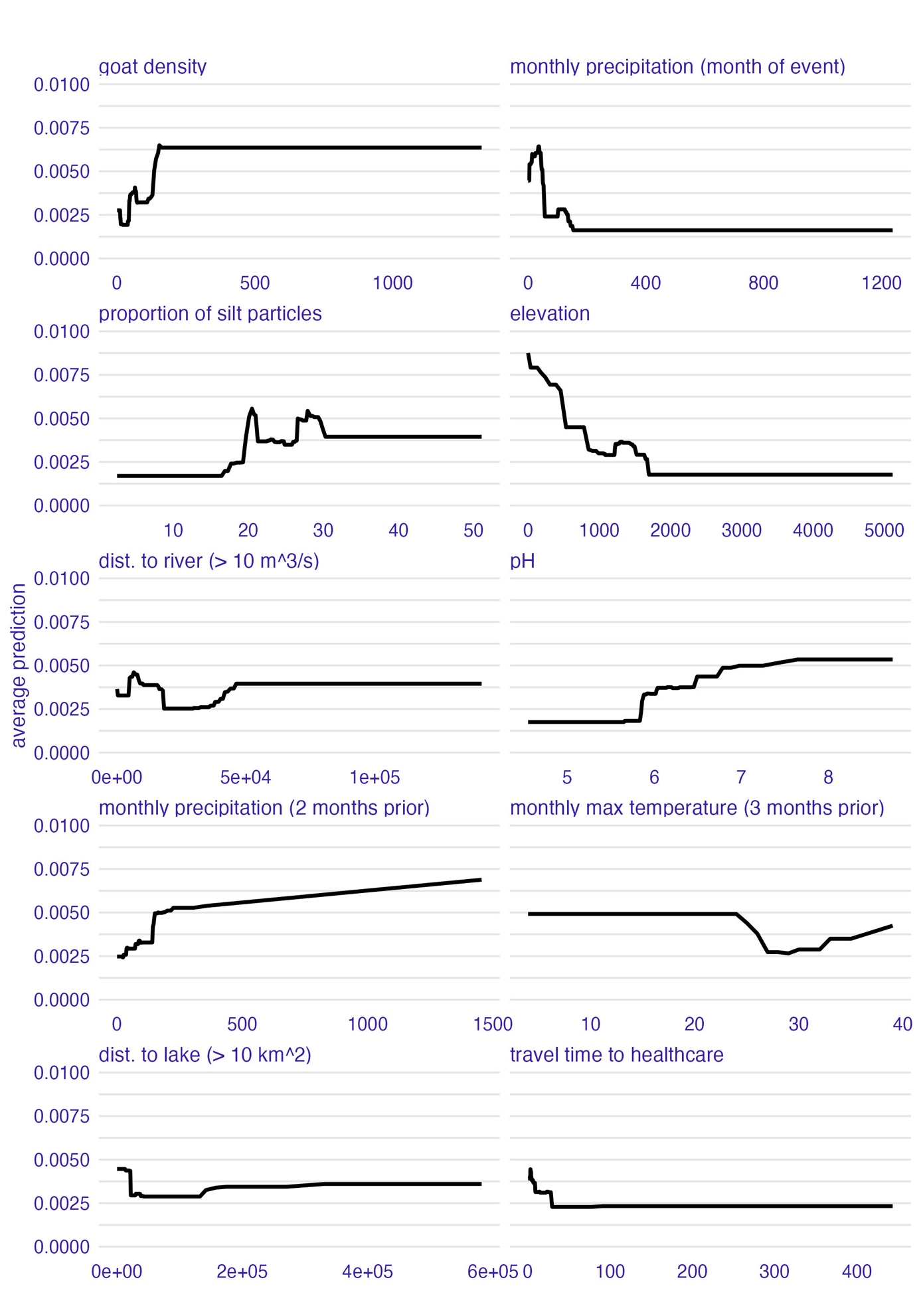

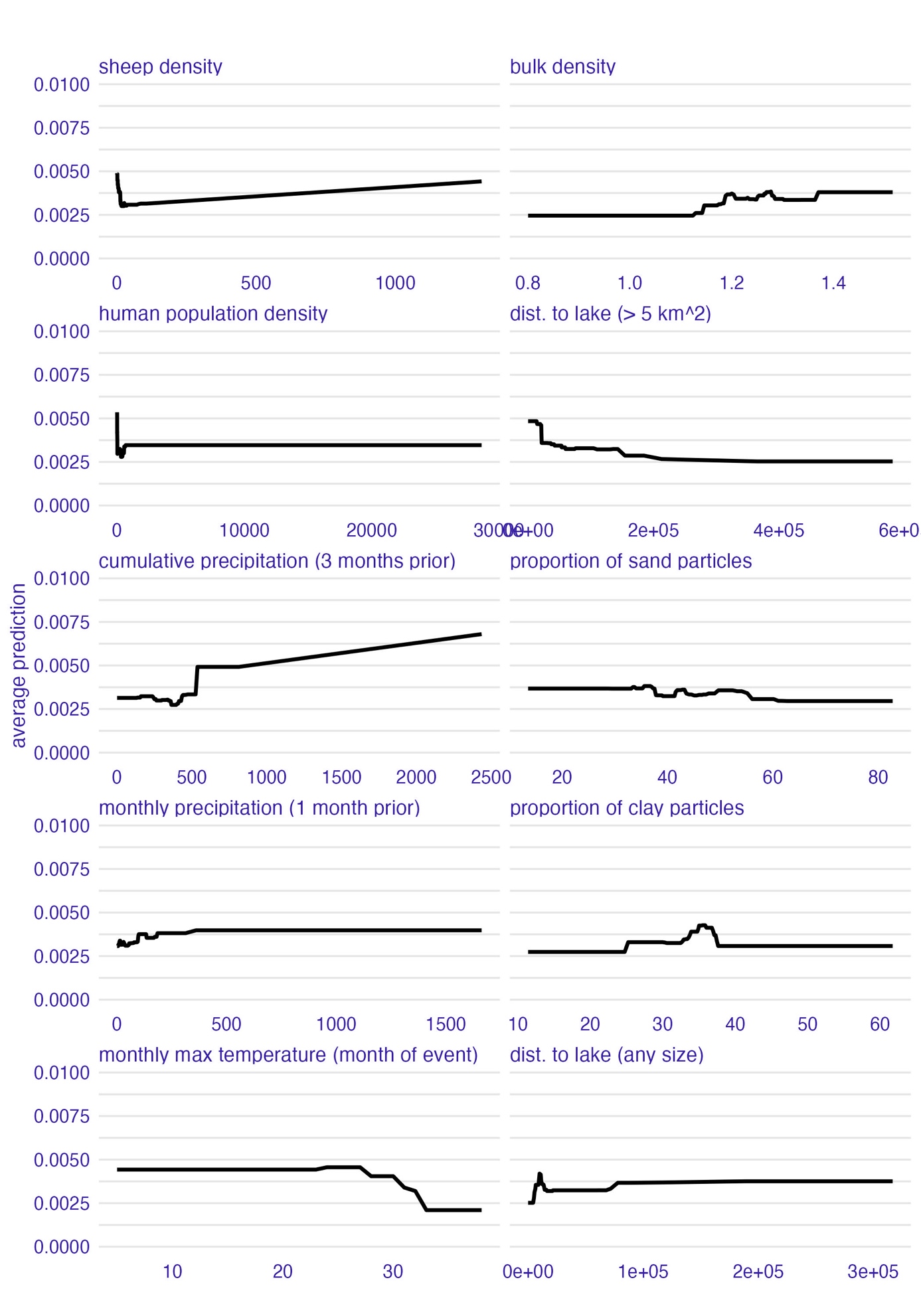

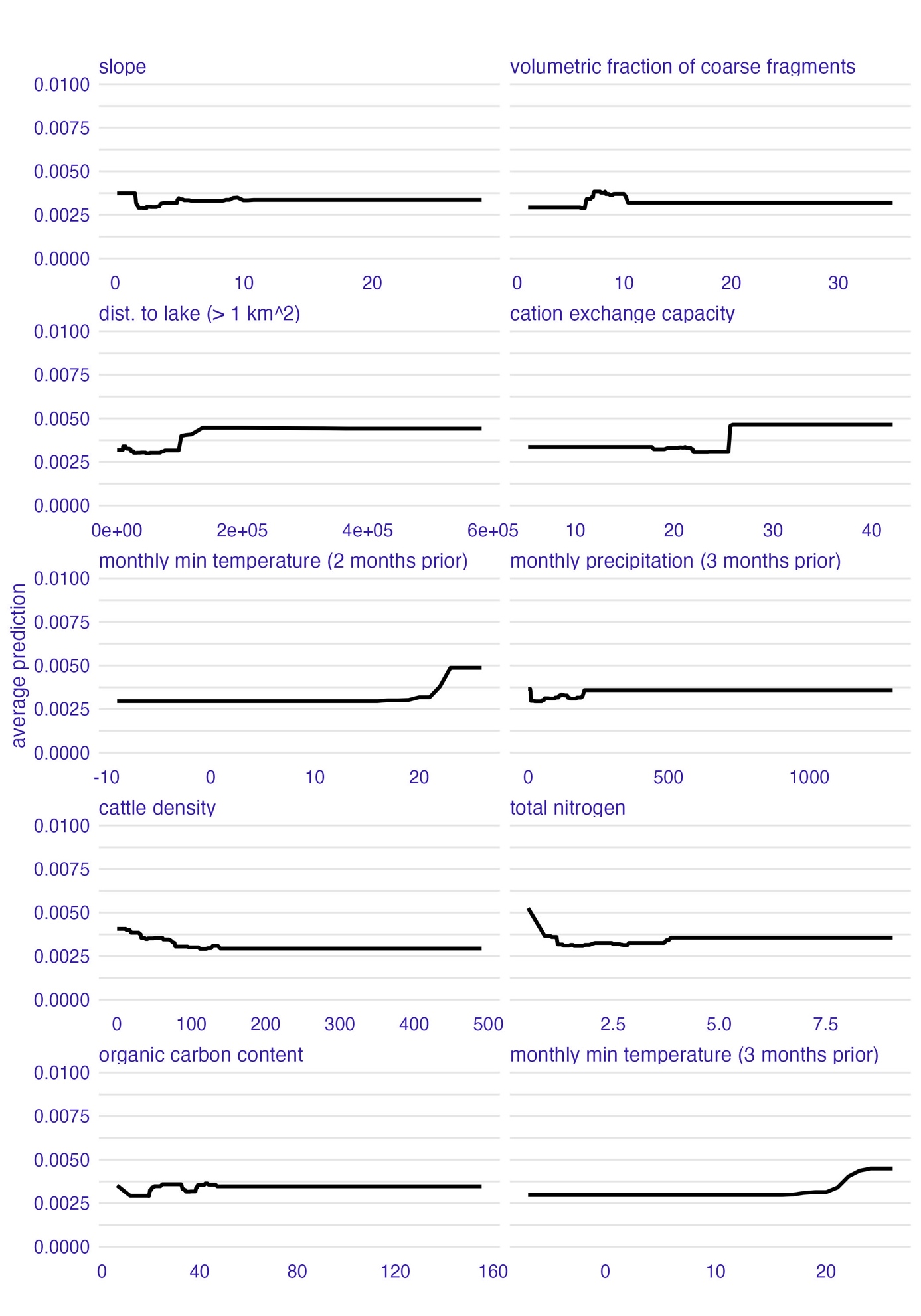

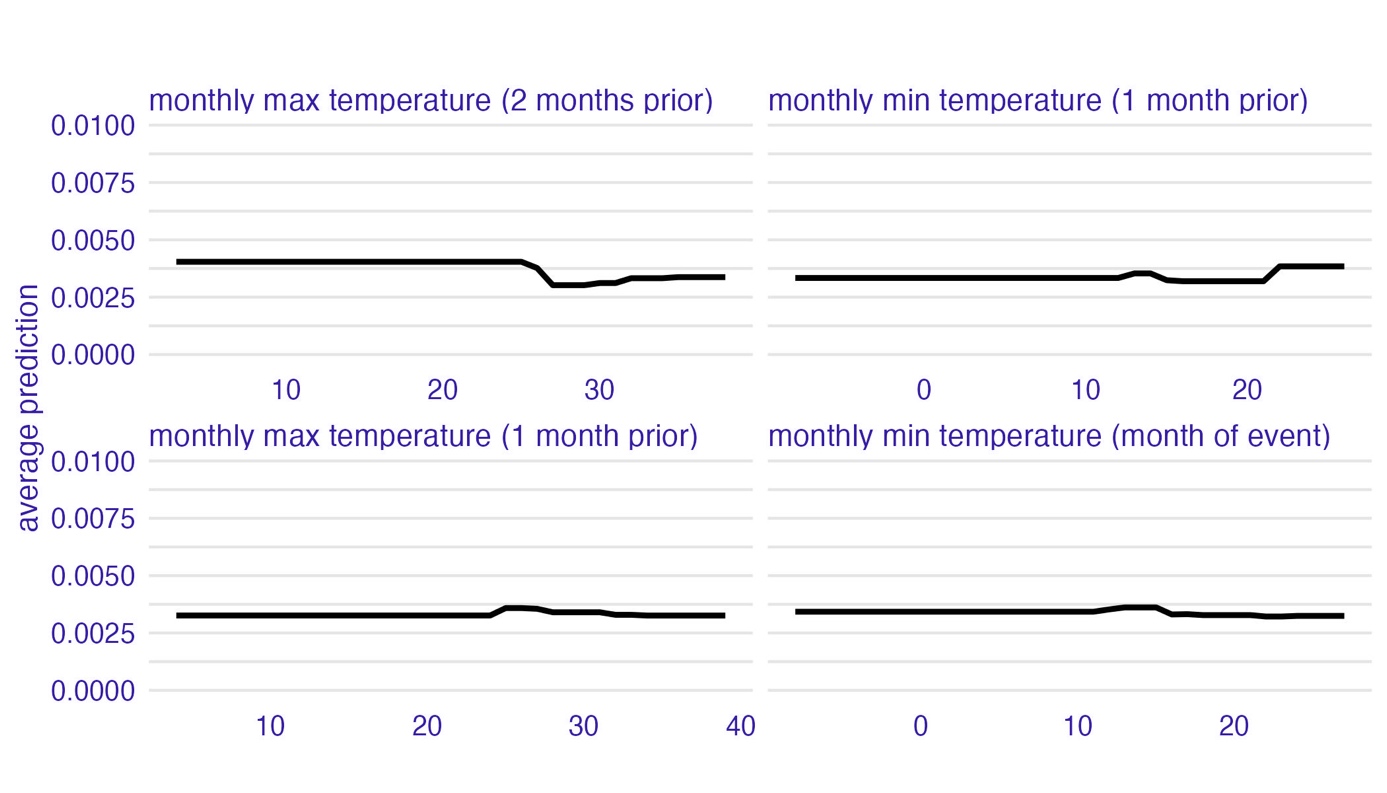

**Figure S4. Partial dependence plots for all 34 predictor variables used in XGBoost model training.**

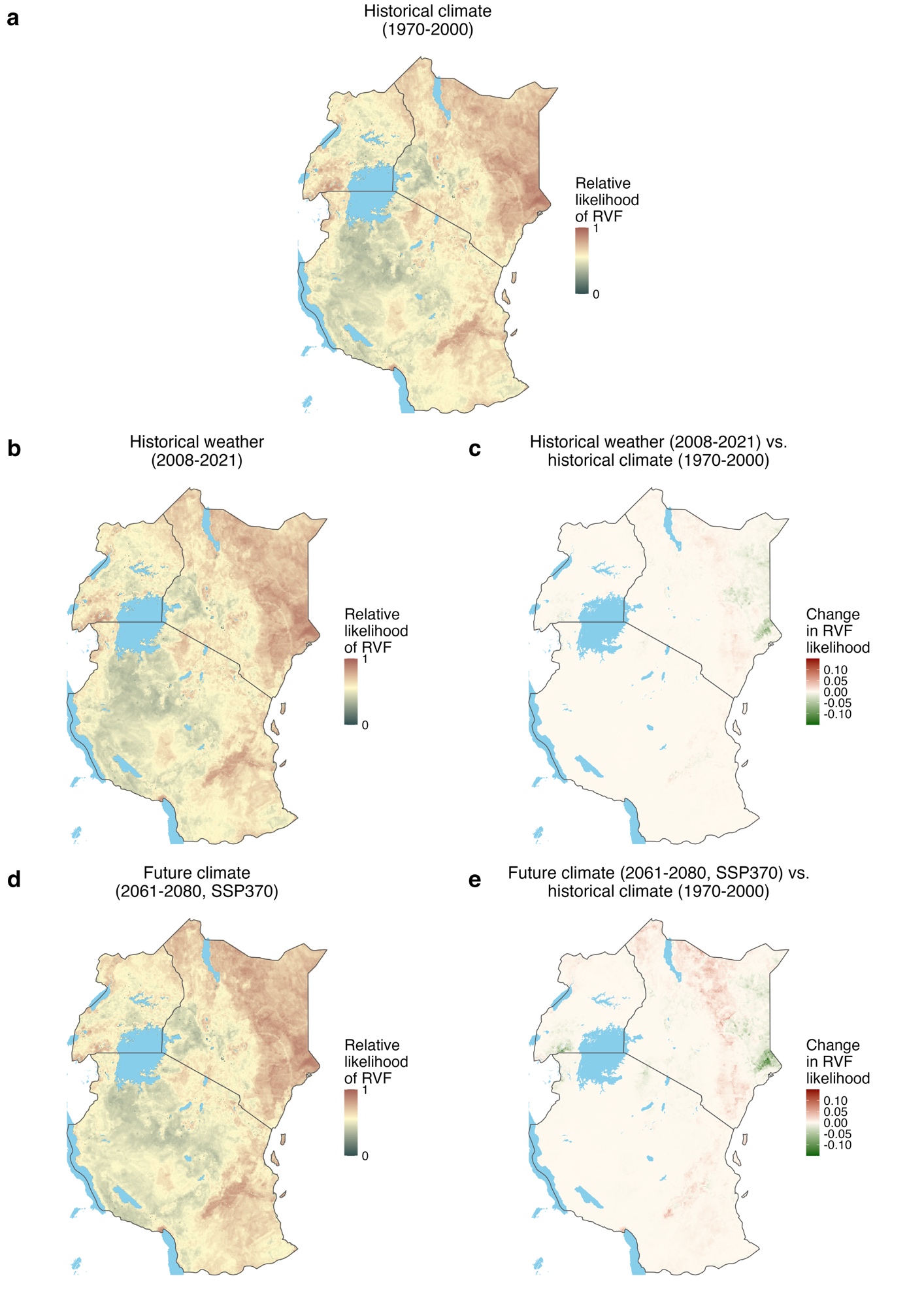
**Figure S5. Predicted relative likelihood of inter-epidemic RVF for different climate and weather scenarios.** Panel **a** shows the average prediction across the 12 calendar months represented in the historical climate data (1970-2000), panel **b** shows the average prediction across all years and calendar months from 2008-2021 (derived using monthly weather data from this time period), and panel **d** shows the average prediction across the 12 calendar months and 11 climate models for the 2061-2080 time period and the SSP370 future climate scenario. We chose to focus on the 2061-80 time period and the SSP370 scenario as they represent the most extreme departure from historical climate that we considered. In panels **a**, **b**, and **d**, the colorbars show predicted relative likelihood of RVF on a log10 scale to help emphasize intermediate values. Panel **c** shows the difference between panels **b** and **a**. Panel **e** shows the difference between panel **d** and panel **a**.

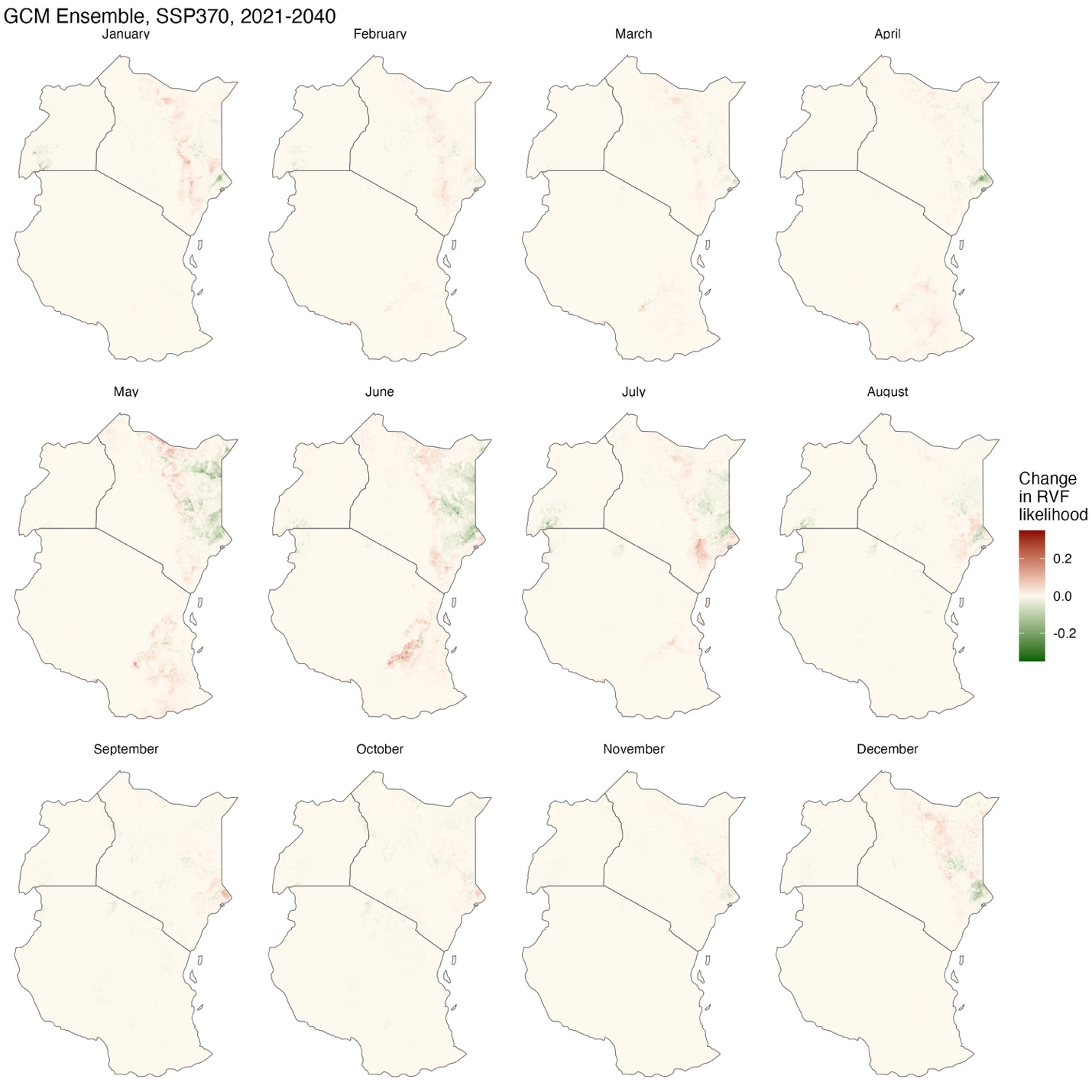

**Figure S6. Difference in monthly predicted inter-epidemic Rift Valley fever risk for 2021-2040 under the SSP370 scenario compared to historical climate (1970-2000).** Ensemble predictions for each month under this future climate scenario were generated by averaging the predictions driven by 11 different climate models. We then took the difference between these ensembled monthly predictions and the monthly predictions generated using historical climate data describing the 1970-2000 timeframe.

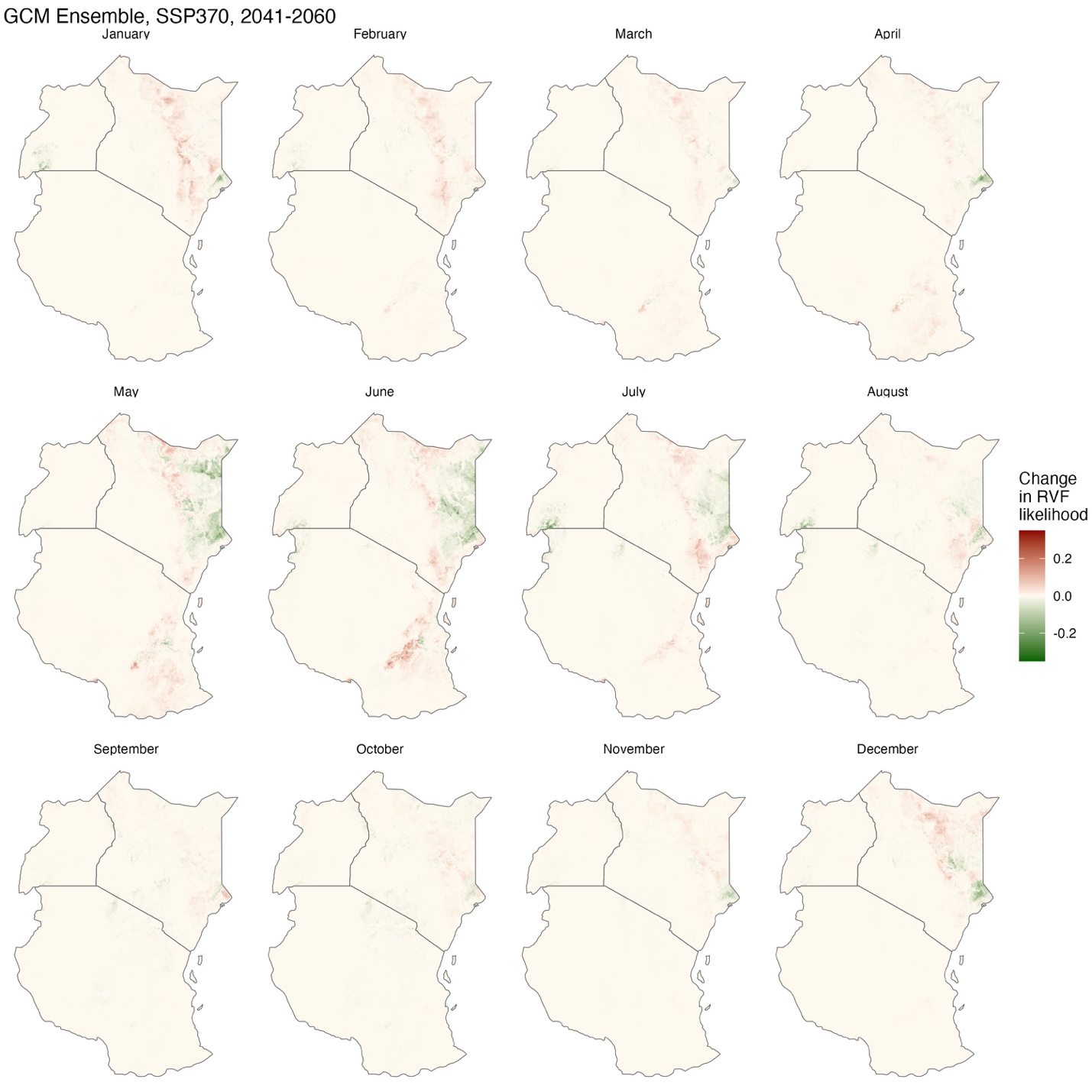

**Figure S7. Difference in monthly predicted inter-epidemic Rift Valley fever risk for 2041-2060 under the SSP370 scenario compared to historical climate (1970-2000).** Ensemble predictions for each month under this future climate scenario were generated by averaging the predictions driven by 11 different climate models. We then took the difference between these ensembled monthly predictions and the monthly predictions generated using historical climate data describing the 1970-2000 timeframe.

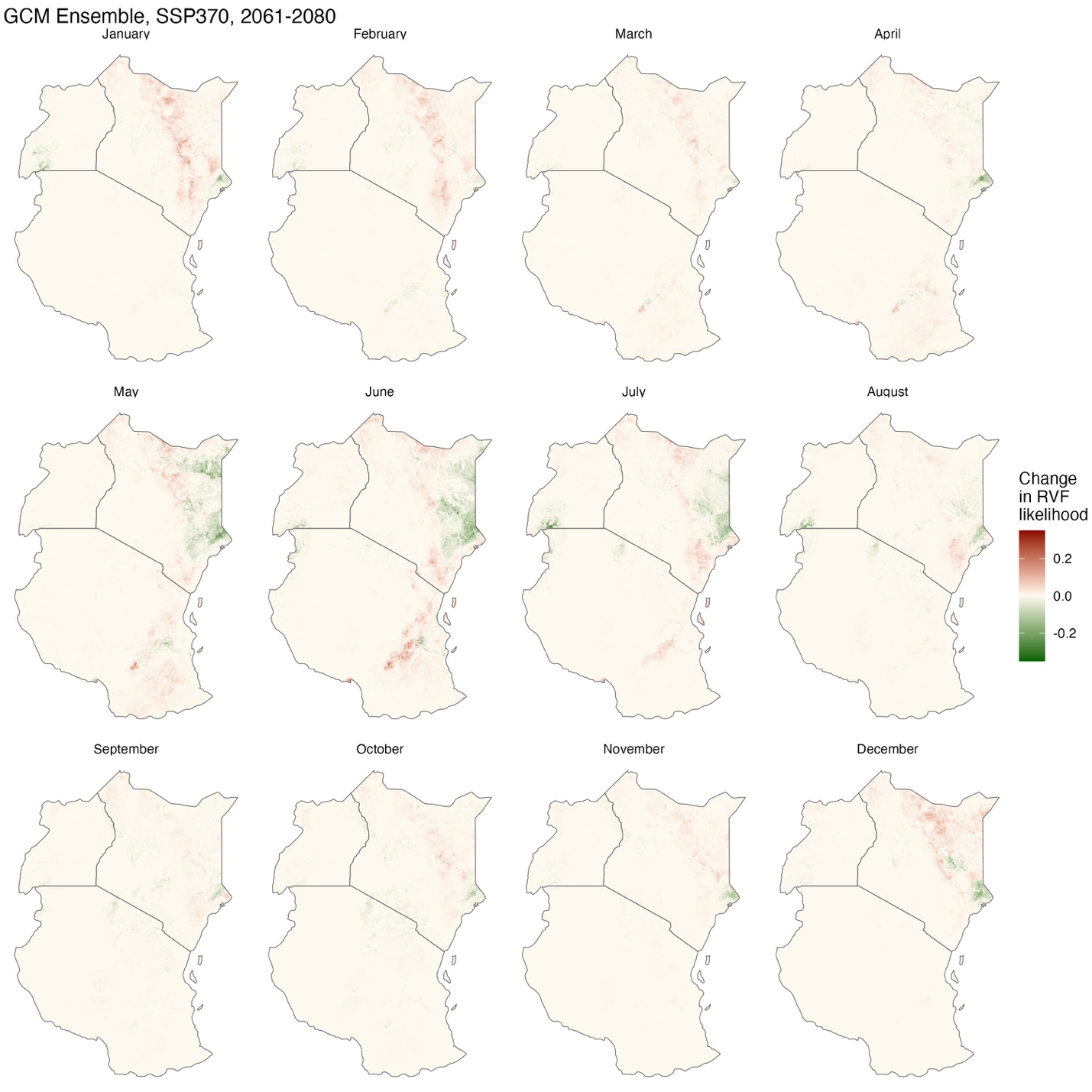

**Figure S8. Difference in monthly predicted inter-epidemic Rift Valley fever risk for 2061-2080 under the SSP370 scenario compared to historical climate (1970-2000).** Ensemble predictions for each month under this future climate scenario were generated by averaging the predictions driven by 11 different climate models. We then took the difference between these ensembled monthly predictions and the monthly predictions generated using historical climate data describing the 1970-2000 timeframe.

**PART 4: REFERENCES**
